# Environmental determinants of *E. coli*, link with the diarrheal diseases, and indication of vulnerability criteria in tropical area (Kapore, Burkina Faso)

**DOI:** 10.1101/2021.04.21.21255867

**Authors:** Elodie Robert, Manuela Grippa, Dayangnéwendé Edwige Nikiema, Laurent Kergoat, Hamidou Koudougou, Yves Auda, Emma Rochelle-Newall

## Abstract

In 2017, diarrheal diseases were responsible for 606 024 deaths in Sub-Saharan Africa. This situation is due to domestic and recreational use of polluted surface waters, deficits in hygiene, access to healthcare and drinking water, and to weak environmental and health monitoring infrastructures.

*Escherichia coli* (*E. coli*) is an indicator for the enteric pathogens that cause many diarrheal diseases. The links between *E. coli*, diarrheal diseases and environmental parameters have not received much attention in West Africa, and few studies have assessed health risks by taking into account hazards and socio-health vulnerabilities. This case study, carried out in Burkina Faso (Bagre Reservoir), aims at filling this knowledge gap by analyzing the environmental variables that play a role in the dynamics of *E. coli*, cases of diarrhea, and by identifying initial criteria of vulnerabilities. A particular focus is given to satellite-derived parameters to assess whether remote sensing can provide a useful tool to assess health hazard.

Samples of surface water were routinely collected to measure *E. coli,* enterococci and suspended particulate matter (SPM) at a monitoring point (Kapore) during one year. In addition, satellite data were used to estimate precipitation, water level, Normalized Difference Vegetation Index (NDVI) and SPM. Monthly epidemiological data for cases of diarrhea from three health centers were also collected and compared with microbiological and environmental data. Finally, semi-structured interviews were carried out to document the use of water resources, contacts with elements of the hydrographic network, health behaviors and conditions, and water and health policy and prevention in order to identify the initial vulnerability criteria.

A positive correlation between *E. coli* and enterococci in surface waters was found indicating that *E. coli* is an acceptable indicator of fecal contamination in this region.

*E. coli* and diarrheal diseases were strongly correlated with monsoonal precipitation, in situ SPM, and Near Infra-Red (NIR) band between March and November. Partial least squares regression showed that *E. coli* concentration was strongly associated with precipitation, Sentinel-2 reflectance in the NIR and SPM, and that the cases of diarrhea were strongly associated with precipitation, NIR, *E. coli*, SPM, and to a lesser extent with NDVI.

Moreover, the use of satellite data alone allowed to reproduce the dynamics of *E. coli*, particularly from February to mid-December (R^²^ = 0.60) and those of cases of diarrhea throughout the year (R^²^ = 0.76). This implies that satellite data could provide an important contribution to water quality monitoring.

Finally, the vulnerability of the population is found to increase during the rainy season due to reduced accessibility to healthcare and drinking water sources and increased use of water of poor quality. At this period, surface water is used because it is close to habitations, free and easy to use irrespective of monetary or political constraints. This vulnerability particularly impacts the Fulani, whose concessions are often close to surface water (river, lake) and far from health centers, a situation aggravated by marginality.

**Author summary:** In 2017, diarrheal diseases were responsible for 1.57 million deaths, principally in Sub-Saharan Africa. Many diarrheal diseases are caused by the presence of enteric pathogens in surface water including *Escherichia coli (E. coli*), a frequently used indicator of the presence of these pathogens. Yet, few studies have been carried out in West Africa to verify this link or to study the relationship between diarrheal diseases, *E. coli* and environmental parameters. These diarrheal diseases also depend on socio-health vulnerabilities. This case study addresses the dynamics of *E. coli* along with another fecal indicator bacteria, enterococci, as well as diarrheal diseases (from three health centers) and socio-health vulnerability (from three villages and Fulani settlements) and their relationship with hydro-meteorological parameters derivable by satellite. The study site is located in the Bagre reservoir in Burkina Faso where Suspended Particulate Matter (SPM) and *E. coli* were monitored over one year. Water was generally polluted by bacteria of fecal origin throughout the year and more so during the rainy season. We observed a significant relationship between *E. coli* and enterococci. *E. coli* concentrations were strongly correlated to, and predicted by, precipitation, satellite reflectance in the NIR band by Sentinel-2, and SPM measured in-situ. Diarrheal diseases were also strongly correlated with these variables as well as *E. coli*. Vulnerability of the population to diarrhea increases during the rainy season. The microbiological health risk is more important during the rainy season, from June to September, and especially concerns the Fulani settlements.

## Introduction

In 2016, waterborne diseases were the second leading cause of death in low-income countries [1]. In particular, in 2016 diarrheal diseases constituted the 8th most important cause of death for all age groups combined and the fifth leading cause of death for children under 5-years [2]. In 2017, diarrheal diseases were responsible for 1.57 million deaths, of which 534 000 were children under 5-years [3]. Five main factors drive this situation: 1) high pollution of surface water by micro-organisms from animal and human faeces, 2) widespread use of untreated surface water for domestic, washing, gardening and recreational uses, 3) deficient hygiene, 4) lack of nearby sanitation and health infrastructures, and 5) weak or non-existent environmental and health monitoring infrastructures [4–5].

The highest health burden occurs in Sub-Saharan Africa where only 56% of the population currently has access to improved water sources^1^ [4]. In this region, in 2017, 85 million people, the large majority of whom live in rural areas, were still using unimproved surface water as their drinking and domestic water source. In addition to this, an estimated 180 million people use unimproved water and a further 135 million people have limited access, i.e. round trip travel time exceeding 30mins, to improved water sources [5]. In the rural areas of Burkina Faso: 35% of the population has access to improved water within 30mins and 33% more than 30mins, with remaining 31% only having access to unimproved water including surface water [5]. However, it is probable that numbers concerning access to improved water by the Joint Monitoring Program (JMP) are overestimated, especially in rural areas. This is due to a combination of socio-economic and environmental factors which may prevent access to improved water [6] or because the continuity of service is not taken into account in the JMP estimations. For example, in Burkina Faso, a bore-hole pump breakdown is officially declared when it lasts at least 1 year and 1 day [7]. It is also worth noting that the coverage of rural water supply services fell by 16 percentage points between 2000 and 2017 [5].

Many diarrheal diseases are caused by the presence of microorganisms in surface water including fecal pathogens: 1) bacteria as typical enteropathogenic *Escherichia coli* (*E. coli*) or enterotoxigenic *E. coli* (gastroenteritis, diarrhea), *Shigella* spp (shigellosis or bacterial dysentery), *Salmonella* spp. (typhoid fever), *Vibrio cholerae* (cholera); 2) parasites as *Cryptosporidium* spp (cryptosporidiosis), *Entamoeba histolytica* (amebiasis or amoebic dysentery); and 3) viruses as rotavirus, norovirus, astrovirus and adenovirus (gastroenteritis, diarrhea). Concerning, the bacterial risk, studies have shown that *E. coli* can be used to indicate a probable presence of fecal borne pathogens of bacterial origin (i.e. enteric pathogens) [8–10]. In temperate areas, improvements in *E. coli* detection methodology made the analysis of thermotolerant (fecal) coliforms unnecessary to assess water quality monitoring [11]. *E. coli* is therefore considered to be the best indicator of fecal contamination or FIB - Fecal Indicator Bacteria [11–13] in these regions. It is recommended by the World Health Organization (WHO) to assess water contamination and the associated risk of diarrheal diseases [14]. However, this indicator has been questioned due to the possibility of naturalization of *E. coli* in the environment [15–18], especially in humid tropical systems rich in nutrients and organic matter (OM) [19].

Few studies have been devoted to *E. coli* in tropical environments, and they mainly focused on coastal zones [20–22], wells, pumps, and boreholes [23–25], or in upland streams in the humid tropics [26–27] but rarely on surface waters in semi-arid areas. Rochelle-Newall et al. [28] underlined the knowledge gaps that remain for tropical ecosystems as regards to *E. coli* survival, transfer and partitioning between free or particle attached fractions [29–30], as well as their relationships with human activities and natural processes and the ultimate impacts on human health. Tropical environments are subject to higher average temperature and more intense precipitation events, which can induce elevated rates of soil erosion resulting in high concentrations of particles in surface waters. This suspended particulate matter (SPM) can provide a refuge from predation for bacteria and create a favorable environment for their development, and multiplication.

Statistically significant relationships between FIB, SPM values and certain environmental parameters (temperature, precipitation) have previously been observed in temperate environments [31–34]. For the humid tropics, the presence of *E. coli* and other FIB in surface waters was found to be linked to temperature, precipitation and onset date of the rainy season, land cover, and sediment resuspension [35–38]. Abia et al. [39], in South Africa, and Cronin et al. [40], in Mozambique, observed an increase in *E. coli* and thermotolerant coliforms respectively, at the onset of the rainy season. Musa et al. [41] also observed a link between *E. coli* and the rainy season in their study from Sudan. Looking at the causes of a diarrhea outbreak in South Africa, Effler et al. [42] showed that the first heavy rains, following 3 months of drought, and the excretion of *E. coli* by cattle were important contributing factors. However, apart from these studies mainly carried out in Southern Africa, and notably in South Africa, important knowledge gaps remain on *E. coli* dynamics in surface water and their links with hydro-meteorological parameters, particularly in West Africa.

Beyond the direct link between the pathogens responsible for diarrheal diseases and environmental parameters, several studies have highlighted the predominance of diarrhea either during the tropical rainy season [43–44], when temperatures are higher [45–47], during floods [48–50], or during periods of abundant precipitation [51, 50] following a dry period. Carlton et al. [52] observed a decrease of -26% in diarrhea cases when the rains followed a wet period. Overall, these studies suggest that diarrhoea rates are positively correlated with increasing temperature and show a more complex relationship with precipitation. Few studies have examined the links between environmental factors and diarrheal diseases in sub-Saharan Africa despite this being the geographic area with the greatest burden. Indeed, Levy et al. [50] found only 11 articles from Sub-Saharan Africa (4 in Southern Africa, 3 in West Africa, 2 in East Africa and 2 reviews) out of 141 produced worldwide. In West Africa, Dunn et al. [53–54] showed that the risk of diarrhea increased in hotter and drier climates, in areas of high rainfall and in low-elevation areas (coastal and along rivers).

Climate change is anticipated to have a strong impact on both the quantity and quality of water resources, potentially boosting the presence, dissemination and transmission of pathogens [55, 52, 56]. Some authors expect that the relative risk of diarrhea in tropical and subtropical regions will increase, on average, by 8-11% for the period 2010-2039, by 15-20% for the period 2040-2069, and by 22-29% for the period 2070-2099 [57]. In addition, the extension of cultivated areas may promote surface runoff which increases SPM and possibly microbial pathogen occurrence in surface waters [58]. Sub-Saharan Africa is undergoing major changes in terms of demographic growth [59] and social and political insecurity that challenge access to clean water and healthcare [60]. Understanding the complexity of interactions between pathogens, environmental parameters, social factors and diarrheal diseases therefore represents an important societal challenge for the forthcoming years.

Water carrying microbiological contaminants represents what can be described as “health hazard”. This health hazard combines with socio-health vulnerabilities of impacted populations to result in a microbiological health risk, in our case, diarrheal diseases. This risk is a multidimensional, multifactorial and constructed process that evolves in time and space [61]. Its analysis requires a holistic approach that integrates biophysical parameters and the characterisation of the pathogens together with the social dimensions. The latter includes exposure (socio-spatial organization and temporal dynamics), sensitivity or susceptibility which refers to social and ecological fragility (practices, water uses, access to drinking water, to a healthcare network, existence of prevention policy, etc.), and forms of resilience [62]. Ultimately, to protect against a risk, a person must perceive the risk as such, know how to protect herself or himself against that risk, and have the means and resources to do so. In addition, inequalities depend on several determinants, in particular, distance [63–64], the size of the household [64] which increases the likelihood of encountering difficulties in paying for care, and the cost of treatment. In Burkina Faso, Haddad et al. [63] showed that socio-economic inequality in access to care is most evident in rural areas: 20% of the richest go to the health center on the first day of illness, whereas 20% of the poorest wait until the 5th day to consult.

Few interdisciplinary studies have combined microbiological monitoring and the study of hydro-meteorological parameters with epidemiological analysis of diarrhea cases and interviews to understand surface water use and healthcare access in tropical areas [65, 26]. To our knowledge, this type of study does not exist in West Africa. Our study applies this original approach to a rural area of Burkina Faso: the Bagre Reservoir region. The objectives of this work were 1) to compare the dynamic of *E. coli* and enterococci in a rural sub-Saharan system, 2) to characterize the hydro-meteorological parameters playing a role in the dynamic of *E. coli*, 3) to reveal the diarrheal epidemiology/incidence in the Kapore region and the associated environmental factors, and 4) to identify the initial criteria of socio-health vulnerabilities.

## Material and methods

### Study Area

The Bagre Reservoir is located in the Center-East region of Burkina Faso (Fig 1A). The average area of the reservoir is 20,000 ha and the maximum area is 25,500 ha, with a volume of 1.7 billion m^3^, corresponding to 14% of freshwater resources in Burkina Faso. The reservoir is located in a basin covering 33.500 km^2^ and belongs to the Nakambe River catchment. The Nakambe catchment is the second largest watershed in Burkina Faso, and it is embedded in the Volta River Basin. The Bagre Reservoir was built in 1992 for hydro-agricultural and hydroelectric purposes. The filling of the reservoir led to numerous modifications in terms of territorial recomposition and the surrounding region has undergone significant changes since the 1980s in terms of health, population growth (natural and migratory), and land use [66–68].

**Fig 1.**
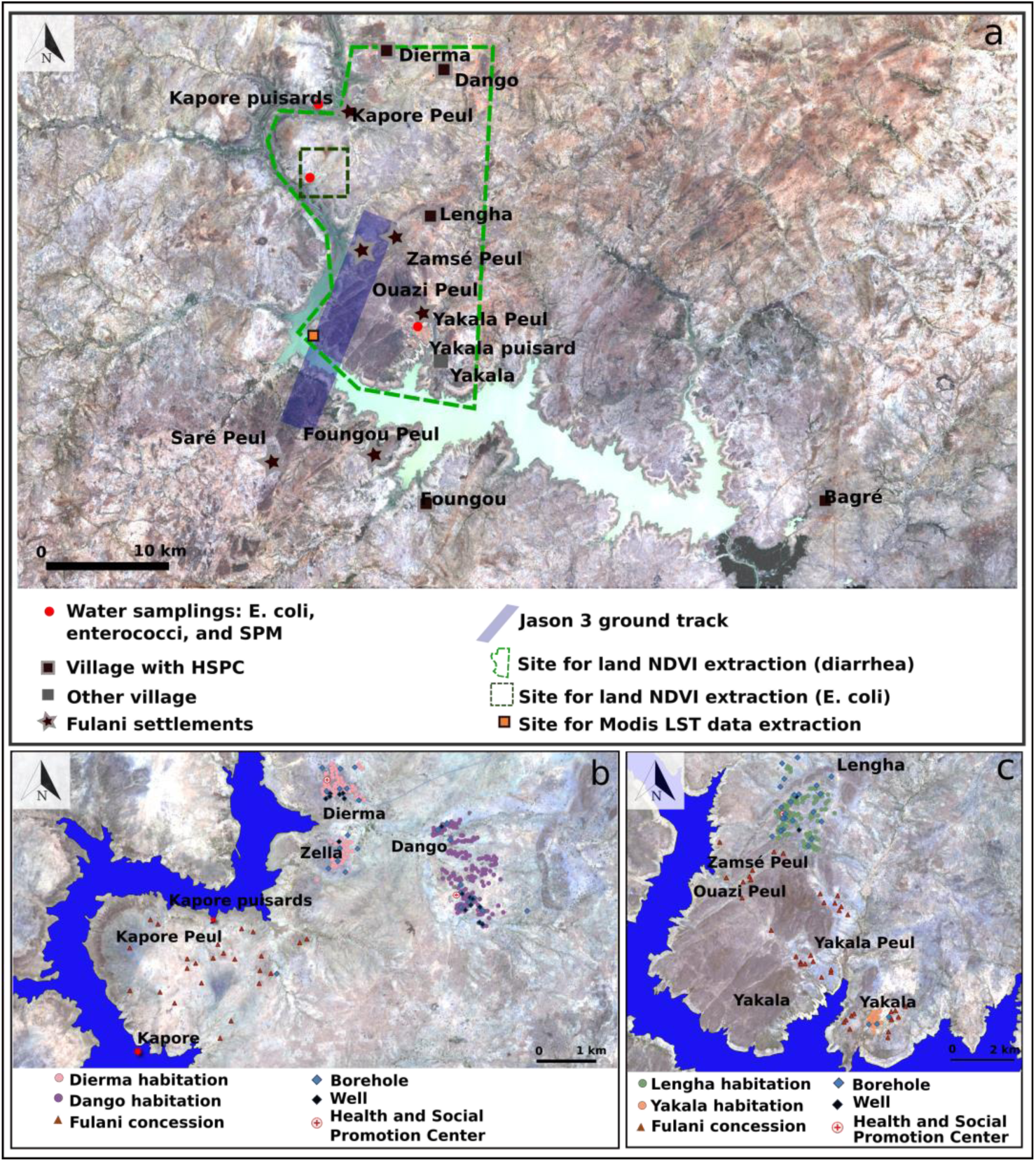
(a) Study Area (Sentinel-2 image 22/04/2018); (b) Zoom on the area of Dango, Dierma and Kapore; (c) Zoom on the area of Lengha, Lengha Peul, Yakala and Yakala Peul.

The region belongs to the Tropical Savannah eco-climatic zone of the Köppen classification. It is characterized by two well marked seasons: a rainy season from June to September, with July, August and September contributing to 67% of the annual rainfall on average, and a dry season from October to May (Fig 2). Between 2000 and 2018, annual precipitation varied between 542.8 mm and 1171.1 mm at the Tenkodogo station with an average of 853.1 mm (Direction Nationale de la Météorologie du Burkina Faso, Fig 2). The annual rainfall in 2017 and 2018 was 645.2 mm and 1057.5 mm, respectively. Thus, 2017 can be considered as a deficit/dry year and 2018 as a humid/wet year. Regarding temperature, the dry season from October to May is characterized by two distinct periods: a dry and cooler period from October to January, and a dry and hot period from February to May. The average temperature varies from 25.6 to 28.4°C (maximum temperature between 33.9 and 37.1°C and minimum temperature between 18.1 and 23.1°C, the minimum being reached in January) during the first, and the average temperature varies between 28.8 to 31.6°C with a maximum average temperature of 33.2°C in April during the second period (Fada N’gourma Station, Direction Nationale de la Météorologie). During the rainy season, average temperatures are close to those observed in December and January (Fig 2) with average temperatures from 26.2 to 29.1°C. Finally, daily surface water temperatures (24-hour average from day and night data) extracted from the MODIS satellite data [69] on an area downstream the sampling site (Fig 1A, see also the satellite data section below) show a seasonal variability similar to air temperature but with a lower seasonal amplitude ranging from 29.7 to 30.3°C (average night temperature from 29.4 to 29.9°C, average day temperature from 30.0 to 30.8°C).

**Fig 2.**
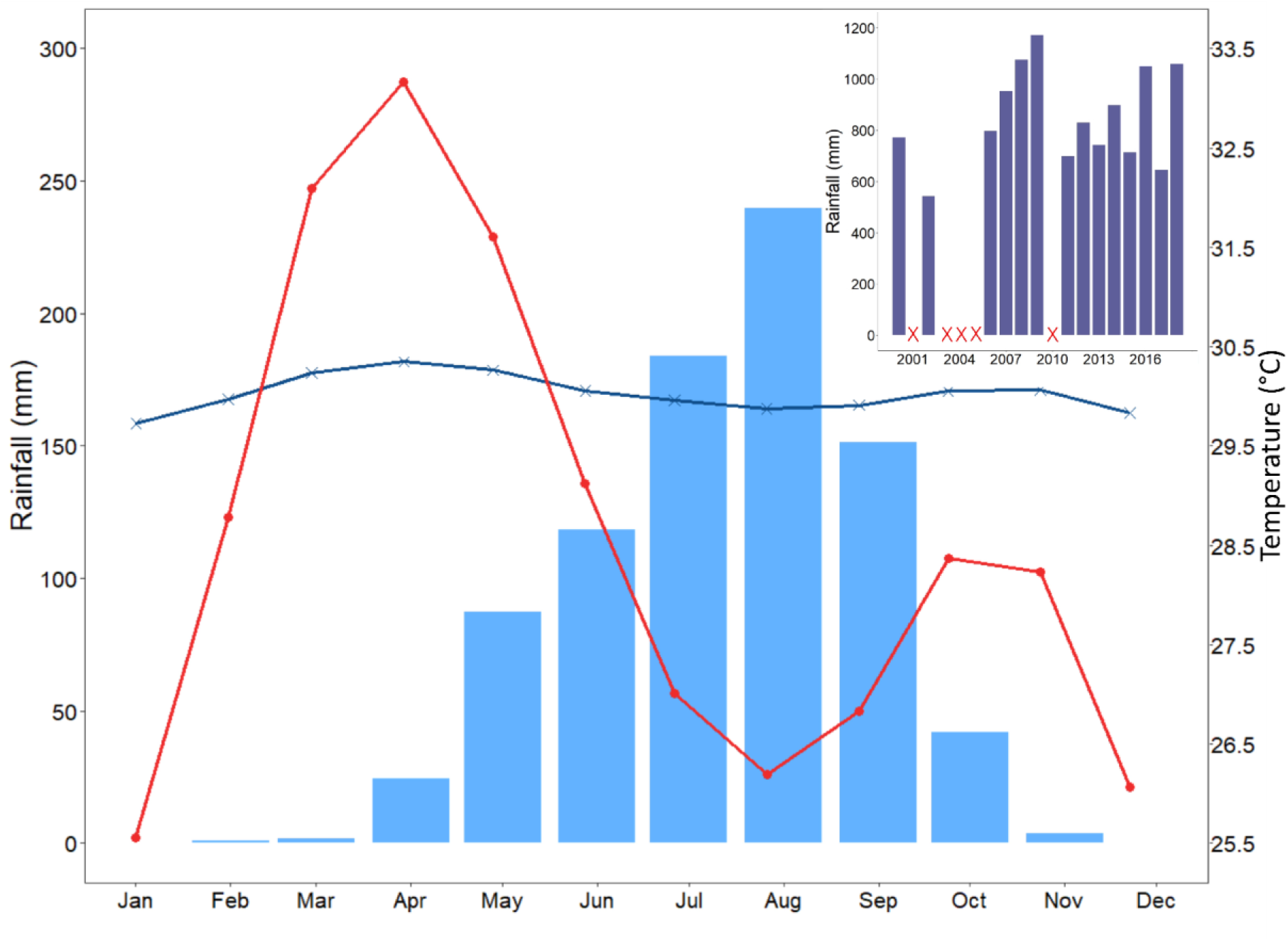
Average monthly rainfall for the period 2000-2018 (bar chart in blue, Tenkodogo station), air temperatures for the period 2000-2018 (red line with dots, Fada N’Gourma station) and satellite_derived surface water temperatures for the period 2000-2018 (blue line with crosses). Annual rainfall from 2000 to 2018 is shown in the inset (no data is available for 2001, 2003-2005 and 2010)

A routine water sampling point was set up in March 2018 at Kapore (11.663010, -0.776650°, Fig 1A) in the upstream part of the Bagre Reservoir to monitor SPM, *E. coli* and enterococci. This site is suitable for satellite monitoring by Sentinel-2 (spatial resolution of 10 - 20 meters) throughout the year. The sample site is located near three farming villages (Dango, Dierma and Lengha), each with a Health and Social Promotion Center (HSPC), and Fulani camps (Fig 1B, C). The main indigenous ethnic group is Bissa, followed by Mossi and Fulani.

The study area is located in the Health district of Garango. The Dierma HSPC was built in 1992, its health area had 4,167 inhabitants in 2018. The Dierma health area includes Fulani concessions near to Kapore (Kapore Peul) which are located between 5.5 and 10 km from the Dierma HSPC and between 2.5 and 7 km from boreholes and wells of Zella (the nearest neighborhood of the village of Dierma, Fig 1B). There is only one borehole at Kapore Peul and some concessions are located more than 3 km away (Fig 1B), forcing people to use surface water including the lake. The Dango HSPC opened in 2006, its health area included 3,999 inhabitants in 2018. Half of the people coming for consultations live outside this health area (e.g. coming from Lengha or Dierma). The village of Dango does not have Fulani settlements (Fig 1B). The HSPC of Lengha was created in 1994 and its health area included 8,396 inhabitants in 2018 (including Koumboré: 2,084 inhabitants, Lengha: 4,243 inhabitants, Ouazi Peul and Zamsé Peul composing Lengha Peul: 746 inhabitants, Massougou: 564 inhabitants, and Yakala: 669 inhabitants). The Zamsé Peul concessions are close to Lengha and people use the nearest Lengha well. The Ouazi Peul concessions are located 2.5 km from the Lengha wells (Fig 1C). The Yakala Peul concessions are located between 3 km and 5.5 km from the first wells of Lengha, 7 km from HSPC and 5 km from the Yakala boreholes (Fig 1C). However, there is a well at Yakala Peul used by nearby concessions but not by those further away (Fig 1C). Finally, Yakala has 3 boreholes and is located 9 km from Lengha (Fig 1C). This situation in the Lengha region forces certain populations, mainly Fulani, to use surface water, including the lake. In addition to the services provided by the HSPC the population also consults traditional healers.

Populations living near the banks of the Bagre Reservoir depend on nearshore water for drinking, cooking, dish and clothes washing, showering or bathing and recreation (Fig 3 A). Land use around the sampling site is dominated by croplands (rain-fed subsistence) with some rangelands, where herds are led to graze and drink. During "normal" rainfall years, the animals gather at the lake in April, when the other water points have dried up, and stay there until June-July. The banks of the lake and the shallow waters are also used for market gardening from March onwards and rice is cultivated during the rainy season. During wet years, the lake water level is sometimes too high to permit market gardening. While 2017 was a particularly dry year (Fig 2), in 2018, the shallow waters and lake banks were used from March onwards for market gardening and the herds arrived earlier than usual, in March (Fig 3 B).

**Fig 3.**
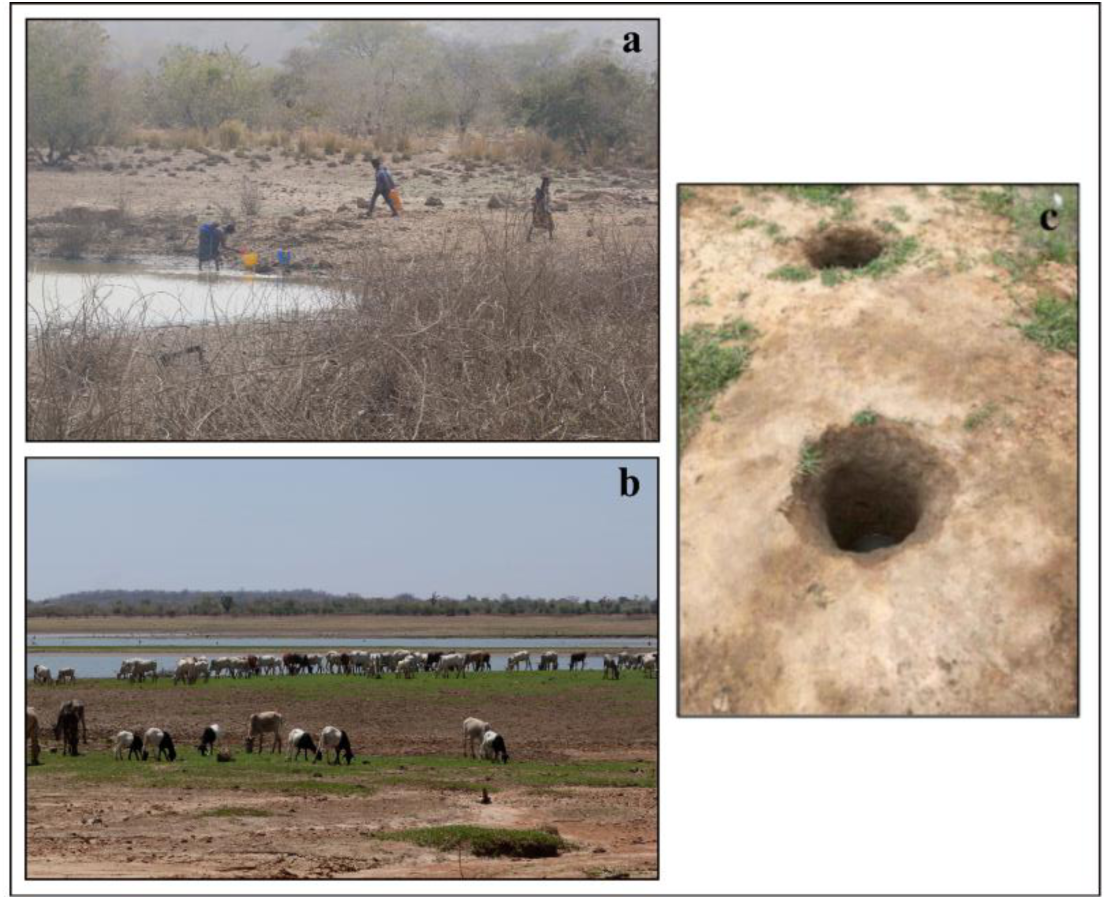
(a) Ouazi Peul: example of site where water of the Bagre Reservoir is drawn for different domestic uses (drinking, cooking, washing, washing clothes), (b) livestock breeding at the Kapore routine site (2018/03/02), (c) “Puisard”.

### In situ data

#### Water sampling

Samples of surface water were routinely collected (just under the surface of water) in clean plastic bottles (500 ml) every 7 days from March 2018 to April 2019 at the Kapore site (Fig 1A) to perform SPM, *E. coli* and enterococci measurements. In addition, water samples were also collected during two field campaigns in March 2018 and July 2018 to measure the same parameters at different sites around the lake. Samples were also collected from “*puisard*s”, open holes dug in the sand of dry riverbeds and banks of the lake where water is drawn for domestic use (Fig 3C).

SPM was measured on duplicate sub-samples (30-100 ml) of water that were filtered on to glass fiber filters (Whatman GF/F 0.7 µm nominal pore size). Filters were pre-weighed after oven-drying at 105°C for 1 h 30 min. After sample filtration, the filters were dried and re-weighed. SPM was calculated as the difference between the dry weights after and before filtering divided by the volume of filtered water.

*E. coli* and enterococci counts were determined using the standardized microplate method (NF EN ISO 9308-3 and NF EN ISO 7899-1 for *E*. *coli* and enterococci counts, respectively). Each sample was incubated at four dilution rates (i.e. 1:2, 1:20, 1:200 and 1:2000 with a volume of 150 µl in well) in a 96-well microplate (MUG/EC and MUD/SF, BIOKAR DIAGNOSTICS) and incubated for 48h at 44°C. Ringers’ lactate solution was used for the dilutions and one plate was used per sample. The number of positive wells for each microplate was noted and the Most Probable Number (MPN) was determined using the Poisson distribution. We used the “MPN CALCULATOR” software to perform the conversion between the number of wells detected and the MPN / 100 ml *E. coli* and enterococci values [71]. The detection limit is 50 MPN / 100 ml. Values below this limit were assigned a value of 1. This method has been used successfully in other tropical environments [27].

#### Epidemiological data

Health data for diarrheal diseases were collected at Garango, which collects all the health data from the health district, with the agreement of the Ministry of Health. This study focused on three HSPC (Dierma, Lengha and Dango, Fig 1B, C), which correspond to the area where the routine measurements were carried out (Fig 1A). Data were collected for the period from January 2013 to March 2019 (except for the Dango HSPC, for which data ends in December 2018). Regarding diarrheal diseases, we considered four groups: typhoid fever, dysentery, non-bloody diarrhea, bloody diarrhea. The data has a monthly time step, except for bloody diarrhea which is listed weekly.

Data from several Annual Action Plans were also collected. They contain information on the area covered by the HSPC (geography, socio-economic and cultural aspects), the distribution of the population across the HSPC area, dominant pathologies, resources (infrastructure, material resources and equipment, means of communication, energy sources, human resources, financial resources), curative activities, and planning of strategic activities for the following year.

### Satellite data

High frequency satellite-derived rainfall data were averaged over the Kapore area, including the two main tributaries located upstream of the sampling point (Fig 1A). We used the GPM IMERGHHV6 data product (30mins), which is suited for monitoring rainfall in tropical regions [72–74]. Rainfall data were cumulated per day and also over 8 day periods to be consistent with the timing of the lake water sampling. Data were collected through the Giovanni website (https://giovanni.gsfc.nasa.gov/giovanni/).

Altimetry data from Jason-3 were used to monitor the water level of the Bagre Reservoir from 01/03/2018 to 01/04/2019. Jason-3 provides 10-day data for the track that crosses the Bagre Reservoir (Fig 1A). They were collected from the Dahiti website [75], https://dahiti.dgfi.tum.de/en/943/water-level-altimetry/).

Sentinel-2 surface reflectance products were used to observe the dynamics of surface water reflectance and area at Kapore. Data were obtained from the THEIA data gateway (theia.cnes.fr). The revisit time is 5 days when Sentinel 2A and 2B are combined. To detect surface water and estimate water areas, we used a threshold on the Modified Normalized Difference Water Index (MNDWI which is calculated as the normalized difference between the green and the SWIR bands). Robert et al. [76] applied a 0.2 threshold for the Agoufou lake in Mali. Environmental conditions for Bagre led us to retain a lower MNDWI threshold, namely 0.075, above which a pixel is considered to be water. Although images are corrected for atmospheric effects using the MAJA software [77–78], we added additional thresholds to avoid undetected residual clouds and retained only blue reflectance values lower than 0.16 and red reflectance values higher than 0.035.

Sentinel-2 Surface reflectance products were also used to monitor the seasonal dynamics of the vegetation close to Kapore by averaging the land Normalized Difference Vegetation Index (NDVI) over two areas: a "small" area which potentially impact the dynamics of *E. coli* at the Kapore site and a "large" area to study the link with the dynamics of diarrhea cases over the whole health area of the three HSPC (Fig 1A). NIR reflectance was used to monitor the dynamics of SPM following Robert al. [79–80, 76].

Finally, the Land Surface Temperature (LST) and Emissivity product MOD11A2 (MODIS sensor on-board TERRA) which provides land surface temperature at 1km resolution, was used to extract the daily surface water temperatures (24-hour average from day and night data). MODIS LST 8-day composite data for daily and nightly overpasses (around 10:30 am and 10:30 pm local time) were extracted using Google Earth Engine (https://earthengine.google.com/). Because of its spatial resolution, LST was extracted over an area slightly downstream from the sample site, where the lake is larger (Fig 1A).

### Interviews

Semi-structured interviews were carried out in March 2016 and 2018 with staff of each HSPC (field health officer and nurse in Lengha, nurse in Dango, field health officer and nurse in Dierma), with a community health club, with 31 local residents from the villages of Dierma and Lengha, as well as with the population of the Fulani settlements (Ouazi Peul, Zamsé Peul, Kapore Peul, Yakala Peul, Lengha Peul, Fig 1B, C). Interviews with health staff focused on: the most important pathologies, diarrheal diseases (incidence, causes, people most affected), use of the health center, accessibility issues, and prevention campaigns. The interview grid for the local residents was developed by adapting the concept of “livelihood” (developed by Chambers [80]) to characterize vulnerability criteria: physical, financial, human and social capital by adding new criteria like practices, distances, seasonality, and marginalities. It was constructed around 4 points and related to: i) activity and environment, use of water resources (type of use - consumption, domestic, recreational, livestock, agriculture - depending on the type resource - improved water source, unimproved water source, river, rain); ii) contacts with elements of the hydrographic network (places and periods); iii) health behaviors and conditions (use of traditional healers, HSPC, self-medication, distance and travel time from HSPC, financial, cultural, and technical availability, etc.); iv) water and health policy and prevention (transformations in access to care following a new building, easier access, prevention and / or awareness campaigns, etc.). Although there were no interviews carried out in Dango, we included cases of diarrhea from this health center, because people from Dierma and Lengha use this health center.

### Ethics statement

The Ethics committee on non-interventional research (CERNI) of the University of Nantes approved the study (approval number: n°19112020). Before starting each interview, we asked for oral consent from each respondent.

### Statistical analysis and software

FIB (*E. coli* and enterococci) data were log_10_ transformed to achieve a normal distribution. We calculated correlations and regressions to identify explanatory variables predicting *E. coli* and cases of diarrhea. First, standard Pearson analysis was performed for correlations between log_10_ transformed *E. coli* and enterococci, and between in-situ SPM and the Near Infrared (NIR) band. Pearson’s correlation (r) is preferred to study these relationships, because a linear relationship can be expected and tested. Spearman’s rank correlation was used for *E. coli* concentrations and environmental parameters (SPM, water level, rainfall, NDVI, number of water pixels) and cases of diarrhea (incidence). Spearman’s rank correlation (r_s_) is preferred in these cases because these datasets do not have normal distributions and linear relations are not expected.

Partial Least Squares (PLS) regression was used to detect dependencies between variables and to construct a predictive model for *E. coli* concentration and for the dynamics of diarrhea cases. We have chosen the PLS regression because it handles missing data by imputation, it is poorly sensitive to multicollinearity, and it decomposes the variability into components (projection in subspaces). PLS regression (for *E. coli* and cases of diarrhea) were applied to obtain a model using all variables, then a model using a smaller number of variables (the main explanatory variables), and last, a model based on key remote sensing variables only. The choice of key variables was based: on the calculation of the percentage variability explained by each variable on the PLS retained components, and the correlation circle. The package CRAN R plsdepot [81] was used for the PLS [82].The CRAN R package ggplot2 [83] was used for plot construction.

## Results

### Water microbiology: bacterial pathogens

From 13 March 2018 to 02 April 2019, 38 measurements of *E. coli* and enterococci were made. The values of *E. coli* vary between less than 50 and 15,000 MPN 100 mL^-1^ and those of enterococci between less than 50 and 26,000 MPN 100 mL^-1^ (Fig 4). 58% of the *E. coli* samples and 70% of enterococci samples exceed USEPA [84] daily bathing water quality standards (single sample: limit of 35 MPN 100 mL^-1^ for enterococci and 126 MPN 100 mL^-1^ for *E. coli*). The majority of samples are above the detection threshold of 50 MPN 100 mL^-1^ so they do not comply with the WHO recommendation for drinking water supply sources that exclude all values above 0 MPN 100 mL^-1^.

**Fig 4.**
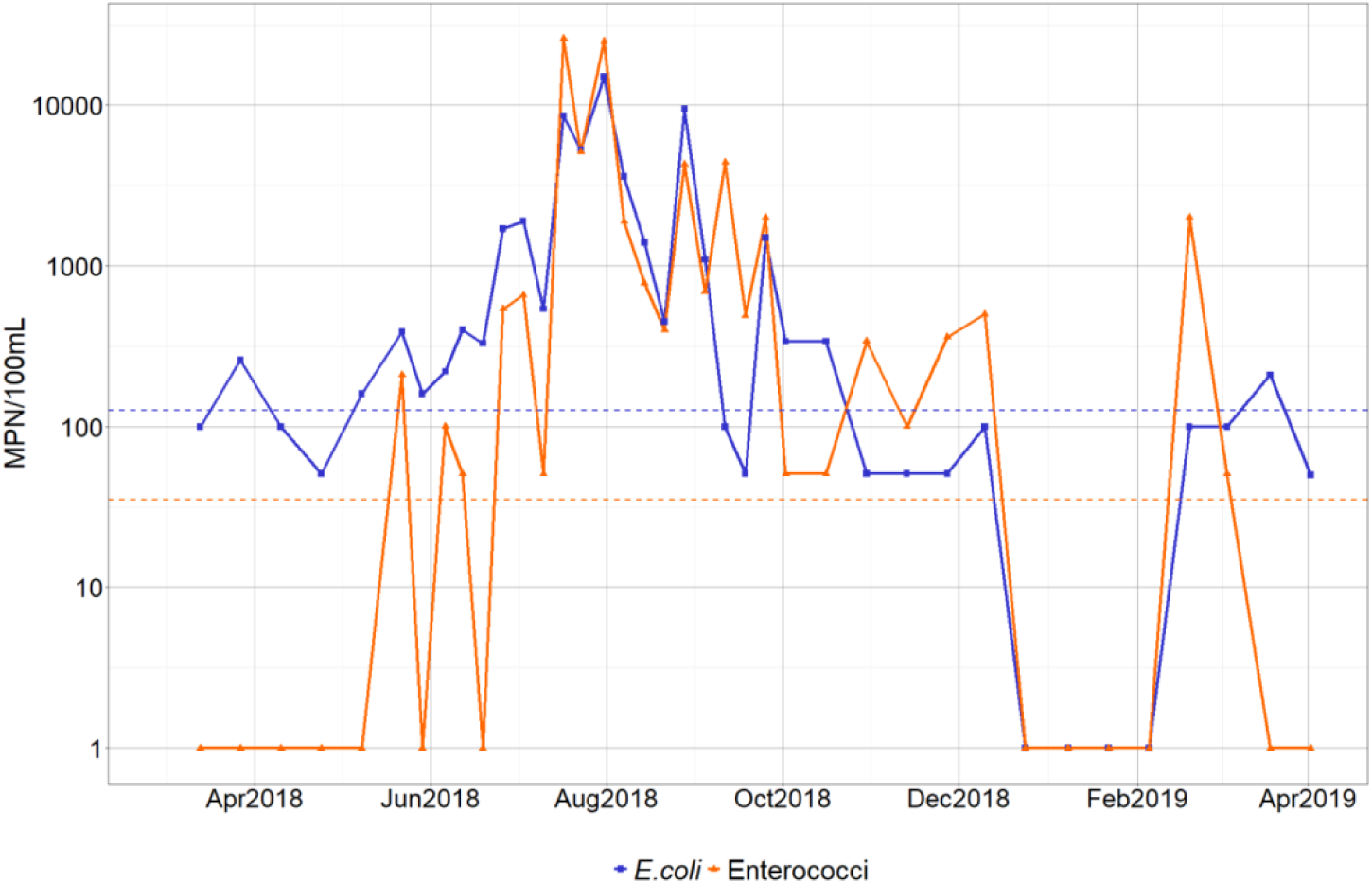
Temporal dynamics of *E. coli* and enterococci at the Kapore site (horizontal dotted lines correspond to USEPA 2012 bathing water quality standard: dotted lines in orange and blue show the limit for enterococci and *E. coli*, respectively)

*E. coli* and enterococci show a similar temporal dynamic (Fig 4). The lowest values are observed from March 2018 to June 2018 and from October 2018 to March 2019. The main peaks are observed in July and at the end of August. Both parameters have very low values from mid-December to early February.

*E. coli* and enterococci numbers are significantly correlated (r = 0.85, without the values of the “puisards”; Fig 5, Table 1). Two specific “outlier” groups can also be noted. The first group, corresponding to the period from March to mid-June (except two dates: 22/05/18 and 06/06/2018) has low values of enterococci (less than 50 MPN 100 mL^-1^) whereas *E. coli* vary between 51 and 330 MPN 100 mL^-1^. The second group, comprised of one value from mid-September and 3 values from November-early December, has relatively low *E. coli* (51 and 100 MPN 100mL^-1^) but high enterococci values (340 to 4400 MPN 100 mL^-1^, Fig 4).

**Fig 5.**
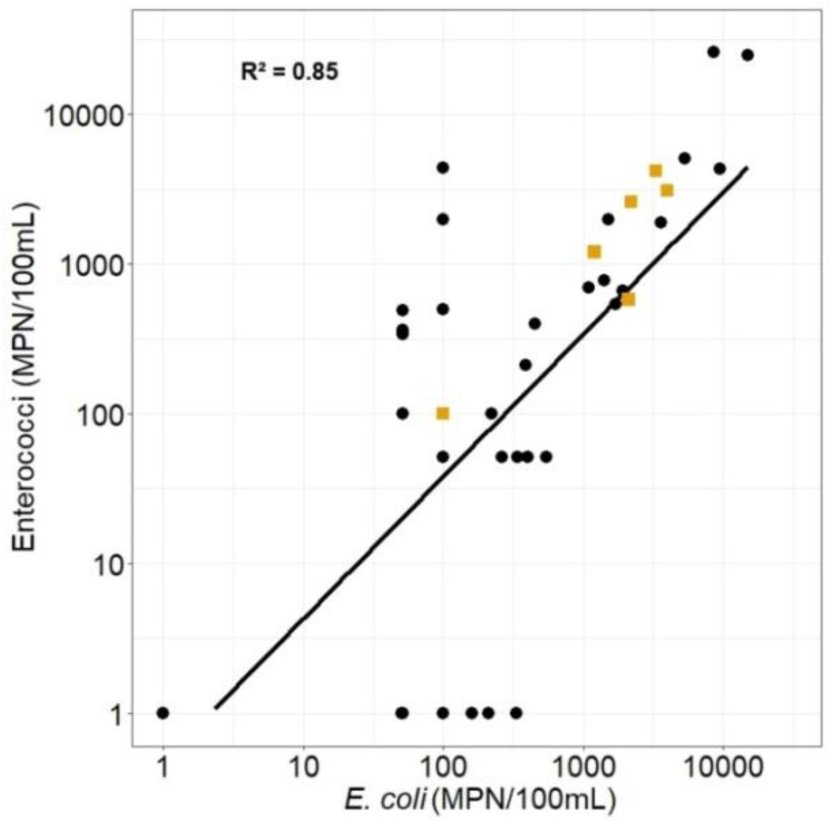
Enterococci vs *E. coli* for the Kapore routine point (N=38), and for “puisards” (N=6, yellow squares).

**Table 1.** Correlation coefficients obtained by regressing *E. coli* and enterococci, and *E. coli* and environmental variables (N=38 for enterococci, SPM, weekly and daily rainfall; N= 16 for cumulative rainfall; N= 22 for land NDVI, Sentinel-2 data; N= 19 for water pixels and water level)

| **Environmental and microbiological variables** | **Correlation** |
| --- | --- |
| **Enterococci** | **$r = 0.85^{****}$** |
| **SPM** | **$r_s = 0.66^{****}$** |
| **Weekly rainfall Kapore** | **$r_s = 0.8^{****}$** |
| **Daily rainfall Kapore** | **$r_s = 0.64^{****}$** |
| **Cumulative rainfall (13/03/2018 to 31/07/2018)** | **$r_s = 0.9^{****}$** |
| **Number of water pixels** | **$r_s = 0.07$** |
| **Water level** | **$r_s = -0.22$** |
| **Surrounding land NDVI** | **$r_s = 0.30$** |
| **Sentinel-2 NIR** | **$r_s = 0.72^{****}$** |

\*\*\*\*corresponds to p-values < 0.0001

The water samples from the “puisards” are consistent with the relationship between *E. coli* and enterococci in the Bagre Reservoir (Fig 5). Values of *E. coli* and enterococci in the “puisards” range from 100 MPN 100 mL^-1^ during the dry season to 3000-4000 MPN 100 mL^-1^ during the rainy season. The values always exceed guideline values (0 MPN 100 mL^-1^) for water for domestic use. The FIB counts observed during the dry season in “puisards” are similar to those observed at the lake routine sampling point. During the rainy season, the values measured in the "puisards" are lower than those measured at the routine point (1200 MPN 100 mL^-1^ versus 5100 MPN 100 mL^-1^).

### Dynamics of *E. coli* in relation to environmental variables

*E. coli* is strongly correlated with precipitation (r_s_ = 0.8 for weekly rainfall, r_s_ = 0.64 for daily rainfall), with SPM (r_s_ = 0.66), and with Sentinel-2 satellite NIR band data (r_s_ = 0.77) (Fig 6, Table 1).

**Fig 6.**
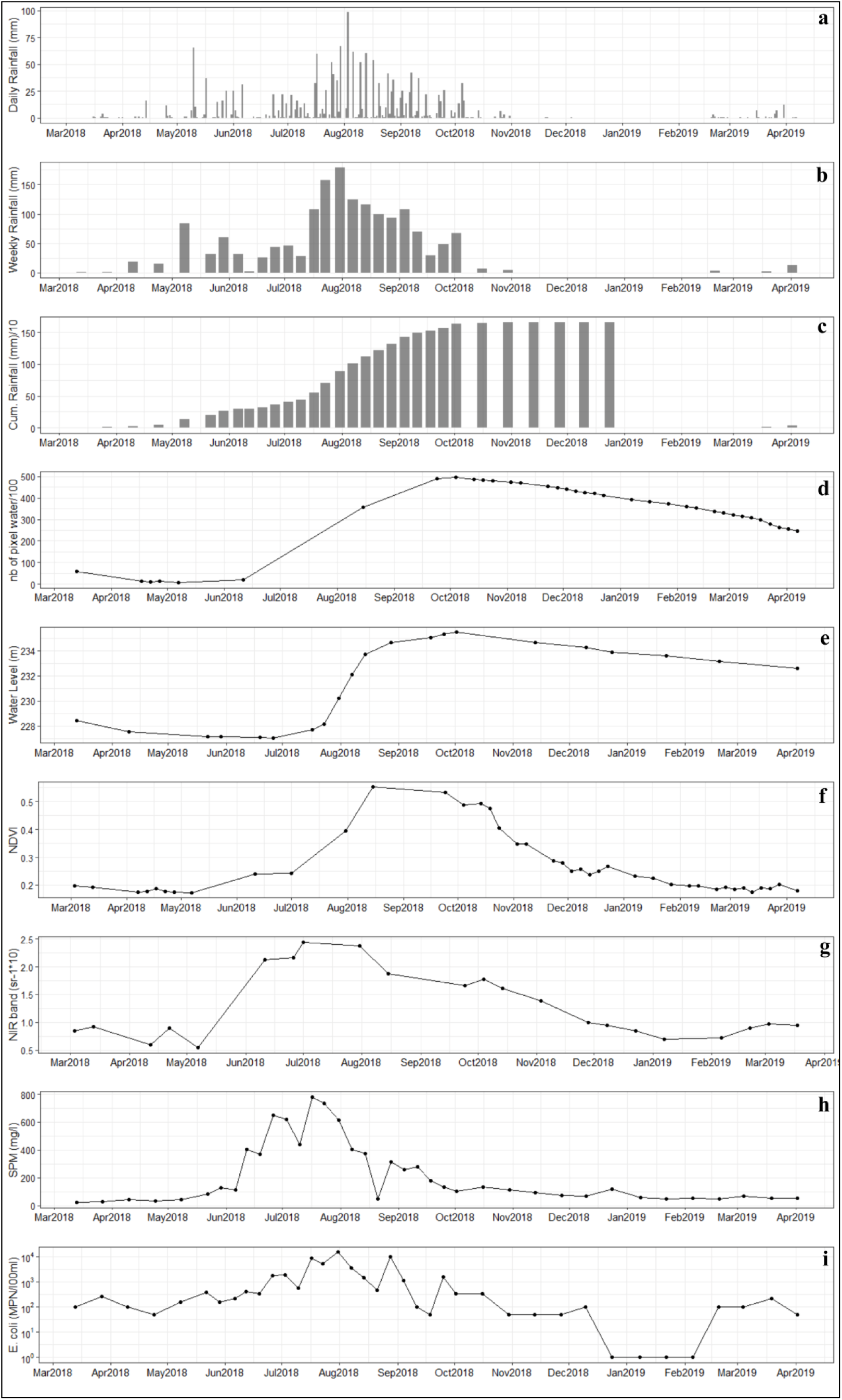
(a) Daily rainfall, (b) Weekly rainfall, (c) Cumulated weekly rainfall, (d) Number of water pixels from Sentinel-2 data, (e) Water level, (f) Average land NDVI from Sentinel-2 data, (g) Water NIR Reflectance (Sentinel-2), (h) SPM, (i) *E. coli*.

Weekly values of SPM and *E. coli* display close seasonal variations (Fig 6H, I). Starting in mid-May, both SPM and *E. coli* values increase in parallel with the arrival of the first heavy rains (Fig 6A) and peak during peak rainy season. Infra-seasonal fluctuations of SPM and *E. coli* during the rainy season seem to correspond to variations in precipitation (Fig 6A, B). In addition, the leading role of precipitation is illustrated by the strong relationship observed between *E. coli* and cumulative rainfall and between SPM and cumulative rainfall from March 13 to July 31, 2018: r_s_ = 0.9, and r_s_= 0.88, respectively (Fig 6C, Table 1). Interestingly, although an increase in bacteria number and SPM concentration is observed when land NDVI is low, both variables decrease when vegetation cover is maximum (September-October) despite significant rainfall (Fig 6F). The lowest values of *E. coli* and SPM are observed from October to early April. In January, in the middle of the dry season, values of less than 51 MPN 100 mL^-1^for *E. coli* and 47.5 to 116.5 mg/l for SPM are found.

A two-week time-lag between the peaks of SPM and *E.coli* is observed at the core of the rainy season with the peak of SPM occurring around mid-July 2018 (780 mg/l) and that of *E. coli* (15,000 MPN 100 mL^-1^) occurring at the end of July 2018.

A good correlation is observed between Sentinel-2 water reflectance and in-situ SPM values, in particular for the Near Infrared (NIR) band (r=0.91), in line with previous studies [85, 75]. Satellite NIR reflectances display a seasonal dynamic similar to that of *E. coli*: increase from May until the end of July-beginning of August, then decrease to reach low values in January, and an increase from February 2019 (Fig 6G).

The first component of PLS regression explains 66% of the variability of *E. coli* from environmental variables and the second one 2% (S1 Fig). The NIR band, SPM, and Weekly rainfall display the highest correlations with the first component of the PLS (i.e. variance explained above 70%, S1 Table, Fig 7). This is in line with results of the one-to-one regressions, although the variable ranking is not in the same. All components have been retained for the *E. coli* predictive model (Fig. 8), resulting in R² equal to 0.66 and RMSE of 1.66 log MPN 100 mL^-1^ (Fig 8A). Using the three main explanatory variables only (S1 Table), R² still reaches 0.54, with a RMSE of 1.81 log MPN 100 mL^-1^ (Fig 8B).

**Fig 7.**
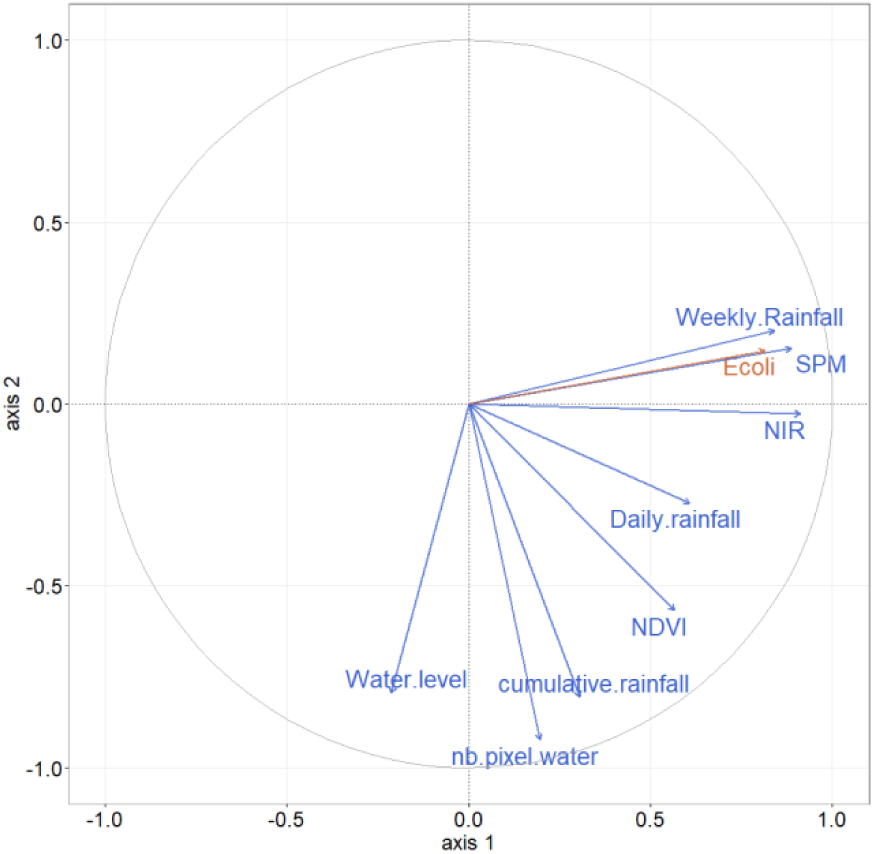
Correlation circle of partial least squares (PLS) regression, illustrating relationships between predictor variables (blue), *E. coli* (orange) and the two first PLS components (first component on the x-axis, second one on the y-axis).

**Fig 8.**
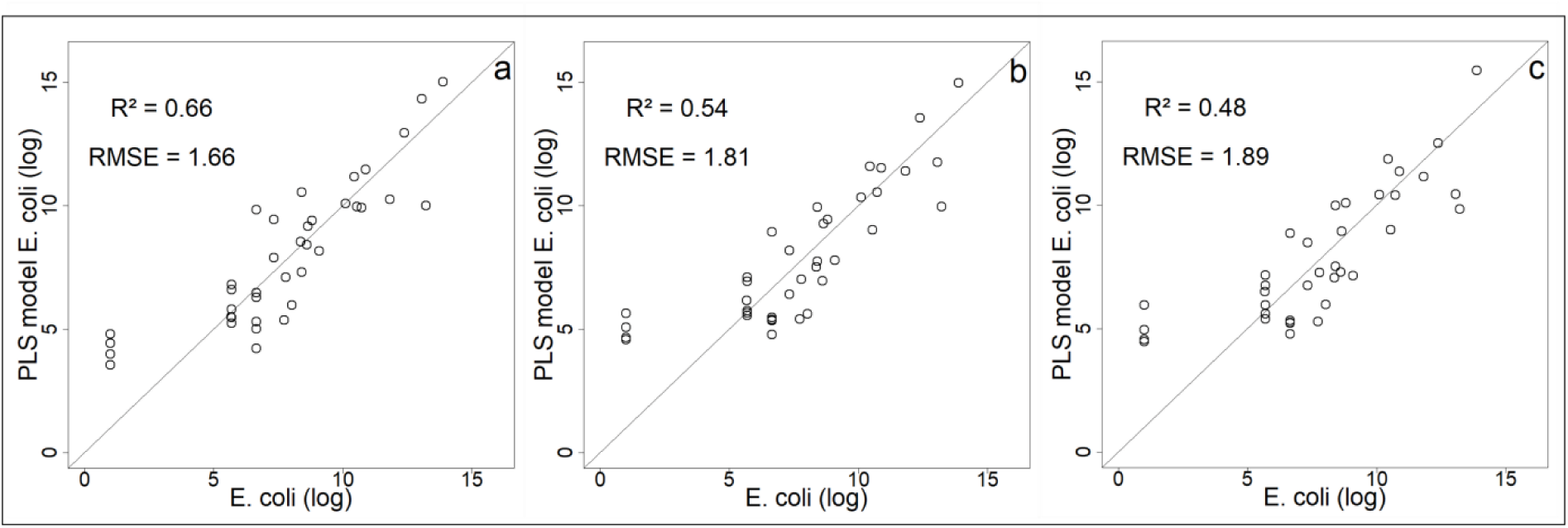
Comparison of models’ predictions with observed *E. coli* (log values) (a) PLS model with all environmental variables and all components; (b) PLS model with NIR band, SPM and Weekly rainfall and all components; (c) PLS model with satellite variables only (NIR band and Weekly rainfall) and all components.

One of our objectives was to question the use of satellite data to monitor the dynamics of *E. coli* to investigate the potential of tele-epidemiological approaches. We therefore chose to perform a PLS regression retaining only the key satellite data (Weekly rainfall and the NIR band, S1 Table, Fig 7) which gave Eq. (1):

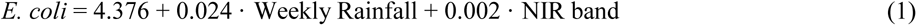

where *E. coli* is expressed in log MPN 100 mL^-1^, Weekly Rainfall in mm, and NIR band in sr*10000. The R² of this model is equal to 0.48 with a RMSE of 1.88 MPN 100 mL^-1^ (Fig 8C).

The model quality limits are linked to individuals 31 to 34 which correspond to the limit of detection of *E. coli* (less than 50 MPN 100mL^-1^ for which it was arbitrarily decided to assign the value "1") (Fig 9). These very low values make the relationship between *E. coli* concentration and the explanatory variables nonlinear. If these data are removed, the quality of the model is improved (S2 Fig), highlighting that our models, whether complete or simplified, represent the dynamics of *E. coli* from February to mid-December (S3 Fig). In addition, it should be noted that if we assign the values of 49, 25, 10, 5 instead of 1 to values below 50 MPN 100mL^-1^, the results are better (R² = 0.8, R² = 0.79, R² = 0.76, R² = 0.74 respectively, and RMSE = 0.98 MPN 100 mL^-1^, RMSE = 1.05 MPN 100 mL^-1^, RMSE=1.21 MPN 100 mL^-1^, RMSE = 1.33 MPN 100 mL^-1^ respectively, S2D, E, F, G Fig). Finally, the models (Fig 9 and S3 Fig) reproduce well the short (weekly) and long term (annual) variability of *E. coli*.

**Fig 9.**
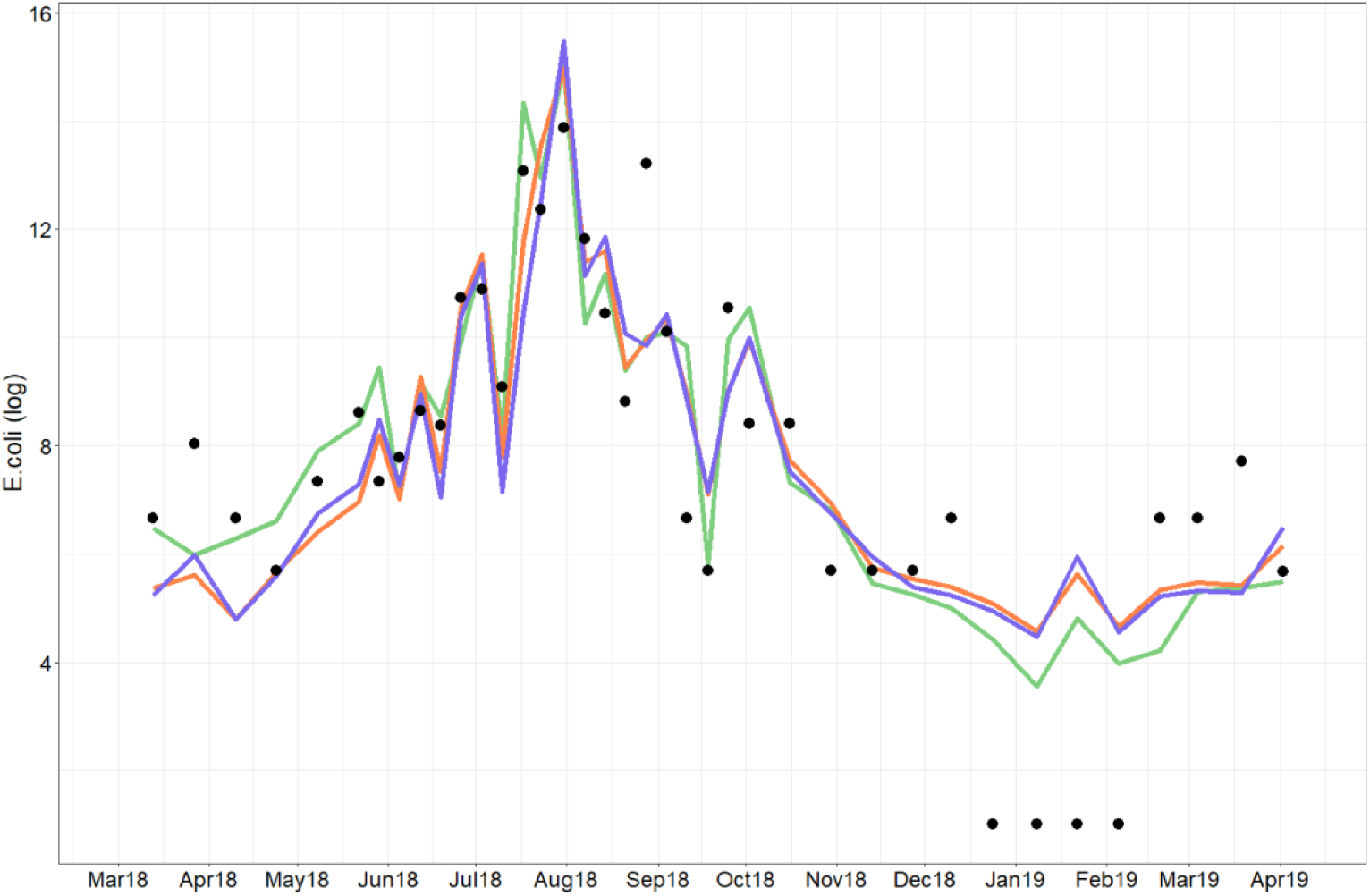
Observed *E. coli* (black point) and predicted *E. coli* (green line for all variables, orange line for NIR band, SPM and Rainfall variables, blue line for satellite variables.

### Analysis of the diarrheal incidence and its temporal dynamics with *E. coli* and environmental variables

In 2018, diarrheal diseases (bloody diarrhea, non-bloody diarrhea, typhoid fever, dysentery) represent 27% of the consultations at HSPC and 11% in medical centers and hospitals in Burkina Faso (2018 statistical directory of Burkina Faso). It is the third cause of consultation. In the Center-East region, these diseases correspond to 6% of the consultations in the HSPC and 1.7% in hospitals.

#### Incidence of diarrheal diseases in the Kapore area

When data from the Dierma, Lengha and Dango HSPCs are pooled together, diarrheal diseases are the third reason for consultation after malaria and respiratory infections. In 2018, children aged 1 to 4 years represented the majority of cases recorded in the 3 HSPC with 39.1% of admissions, followed by children under one year (27.8%), then female adults (14.3%), children aged 5 to 14 years (12.2%), and finally male adults (6.6%). More precisely, at the health centers of Dierma and Dango, adults consulted the most, then children from 1 year to 4 years of age, then those from 5 to 14 years of age, and finally those under the age of 1 year. At the HSPC of Lengha, children aged 1 to 4 years represented the most of the consultations followed by adults, in particular women, children aged between 5 and 14 years, and children under the age of 1 year.

In 2018, non-bloody diarrhea (NBD) disease and dysentery constituted the majority of admissions with 62.3% and 34.4% respectively. There were a few cases of typhoid fever and bloody diarrhea. Whereas the HPSC of Dango reported mainly cases of dysentery, the HPSC of Lengha reported mainly cases of NBD disease as well as cases of dysentery. Similarly, the HPSC of Dierma reported mainly cases of NBD disease, although fewer than those recorded in Lengha. When the data from the 3 HPSCs for 2018 was pooled, a first peak was observed in February, this was followed by a decreasing trend in March and April that thereafter increased to reach a maximum in August after which the values decreased to a minimum in December-January (Fig 10). When observed individually by site, the data from the HPSC of Dango differed slightly from this bulk trend: the first peak occurs in April instead of February, and its minimum is in October. Finally, a time lag between the start of the increase and the main peak differed between HPSC: May and June-August for Dierma, June and August-September for Lengha, June and August for Dango.

**Fig 10.**
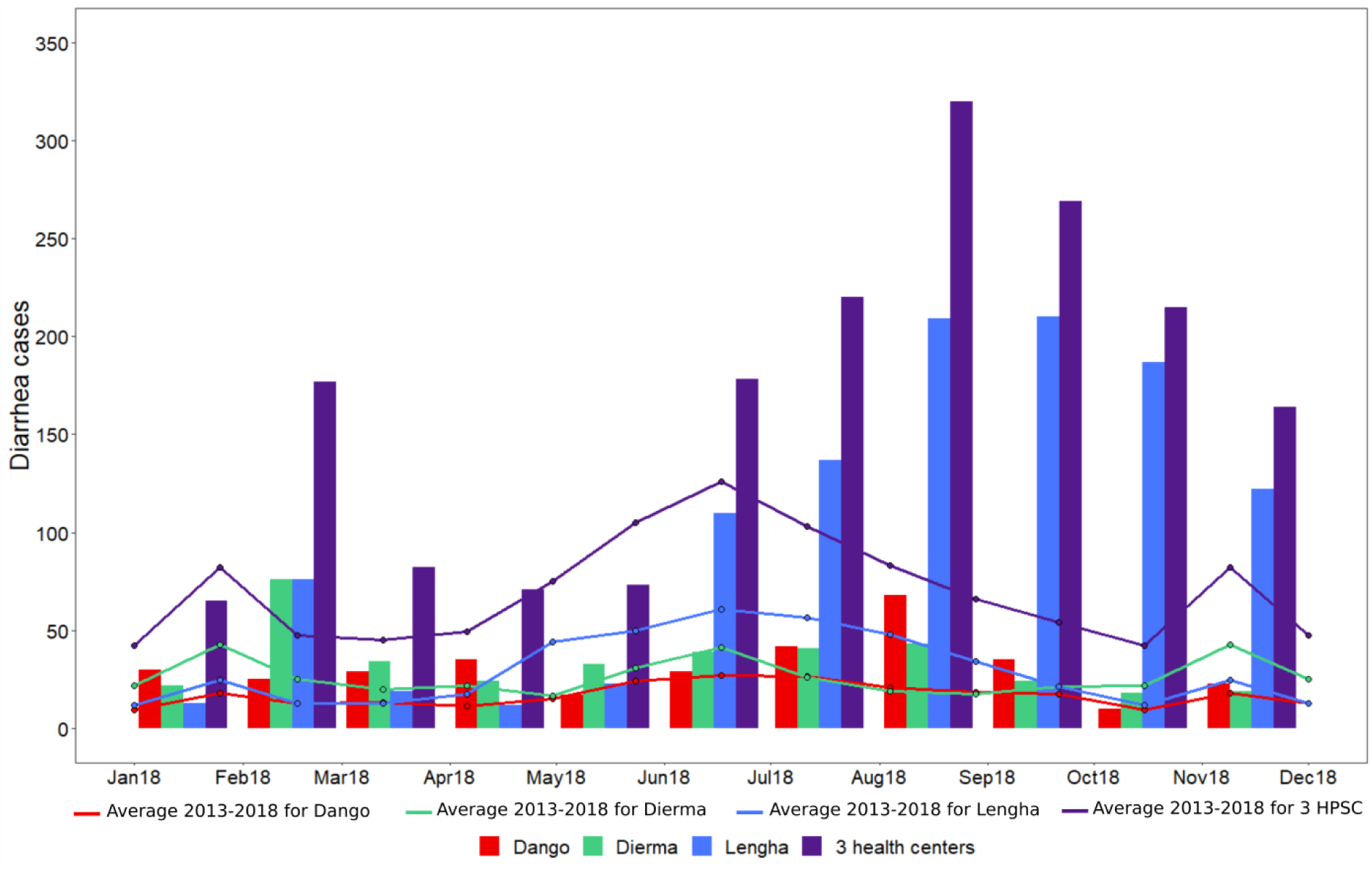
Monthly incidence recorded in 2018-2019 and average recorded for the period 2013-2018 for the 3 HPSC (Dango, Dierma, Lengha) and for all of these 3 HPSC (for January-March 2019 only the data of the HPSC of Dierma and Lengha were available)

For the period 2013-2018, NBD and dysentery constituted the majority of admissions (57.7% and 33.5% respectively) followed by typhoid fever and a few cases of bloody diarrhea. These last two types of disease mainly concern Dierma.

The monthly average established for the period 2013-2018 reveals similar seasonal dynamics. However, the number of cases was much higher in 2018, particularly for Lengha. Indeed, 2018 appears to be an exceptional year in terms of the number of cases recorded compared to other years, especially for the period from June to November and in February.

The temporal dynamic over the period 2013-2018 is similar to that of 2018, although some specificities can be observed. The average cycle of diarrhea cases for the period 2013-2018 of the HPSC of Dango presents a less marked peak in August unlike 2018. Unlike 2018, the second peak is observed in February rather than April, and the minimum is not in October but in January. For Dierma, for the period 2013-2018 the minimum during the first half of the year is observed in June and the maximum in August, whereas in 2018 they are in April and from June to August, respectively. Finally, there are seasonal differences in the use of the health center: consultations are higher in Lengha during the rainy season (56.2%) in contrast to Dango (43.3%) and Dierma (37.7%).

For the period 2008-2018, when we report the number of cases of diarrhea in relation to the total population of each health area (S5 Fig), the numbers for Dierma are higher compared to Dango and Lengha, particularly during the dry season from January to May and to a lesser extent in August. Finally, for the year 2018-2019, Dango and Dierma have higher rates than Lengha from January 2018 to May 2018, in December 2018 and in February and March 2019, while Lengha has higher rates from June to November 2018 (S5 Fig).

#### Relationship between diarrhea incidence and environmental variables and E. coli

The numbers *E. coli* and cases of diarrhea were significantly correlated over the period March 2018 – March 2019 (r_s_ = 0.70, Table 2). There was also a significant correlation between diarrhea cases and SPM values (r_s_ = 0.72, Table 2) with a one-month lag between the peak of SPM in July and the peak of diarrhea cases in August. We also observed a significant correlation between diarrheal cases and precipitation (r_s_ = 0.71, Table 2) and diarrheal cases and surrounding land NDVI (r = 0.68, Table 2).

**Table 2.** Correlation coefficients obtained by regressing diarrhea incidence and *E. coli*, and diarrhea incidence and environmental variables over March 2018-March 2019.

| Environmental and microbiological variables | Correlation |
| --- | --- |
| *E. coli* | $r_s = 0.70^{**}$ |
| SPM | $r_s = 0.72^{**}$ |
| Rainfall Kapore | $r_s = 0.71^{**}$ |
| Surrounding land NDVI (wide area) | $r_s = 0.68^{**}$ |
| Number of water pixels | $r_s = 0.07$ |
| Water level | $r_s = 0.23$ |
| Sentinel-2 NIR | $r_s = 0.63^*$ |

\*corresponds to p-values < 0.05
\*\*corresponds to p-values < 0.01

The seasonal dynamics were similar between April and December. A one-month lag between the peaks of NIR band and cases of diarrhea (r_s_ = 0.63), occurring in July and August respectively, was observed similar to that observed for in-situ SPM (Fig 11).

**Fig 11.**
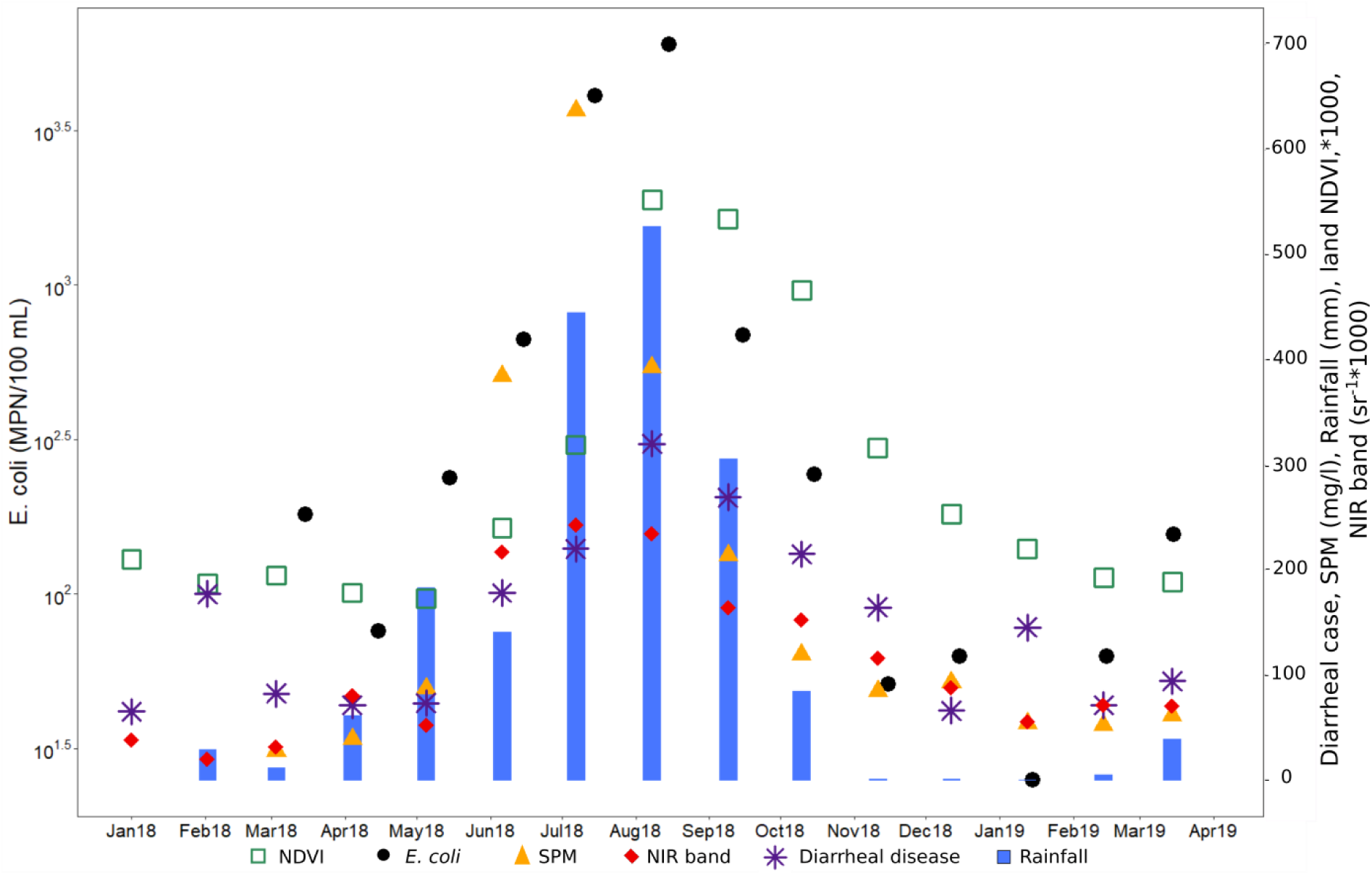
Monthly average of SPM (mg / l), *E. coli* (MPN / 100mL), NIR band (sr^-1^*1000) and land NDVI (* 1000), rainfall (mm), and cases of diarrhea for all 3 HPSC (Dierma, Lengha and Dango) over March 2018 - March 2019.

The first component of the PLS regression explains 78% of the variability of diarrhea cases and the second one 2% (S4 Fig). Monthly rainfall, NIR band and *E. coli* are the main explanatory variables of the first PLS component (explained variance above 70%, S2 Table, Fig 12), and to a lesser extent the SPM and NDVI (explained variance equal to 68% and 67% respectively, S2 Table, Fig 12). Compared to the results of the one-to-one regressions, the PLS shows that the NIR band variable has a more important role than SPM, and that the NDVI has a less important role. We decided to retain these 5 variables as the main explanatory variable to set up the simplified model. As done for the *E. coli* analysis, all components have been retained for the predictive models. When all the variables are retained, the R² of the model is equal to 0.81 with an RMSE of 32.08 cases of diarrhea (Fig 13A and Fig 14). When only the main explanatory variables are used (Monthly rainfall, NIR Band, *E. coli,* SPM, and NDVI), the model presents an R² = 0.82 with an RMSE of 31.00 cases of diarrhea (Fig 13B and Fig 14). The use of satellite data that best explain PLS component 1 variance (S2 Table, Fig 12) provides an R² = 0.76 and an RMSE = 35.14 (Fig 13C and Fig 14, Eq. (2)) and the following model:

**Fig 12.**
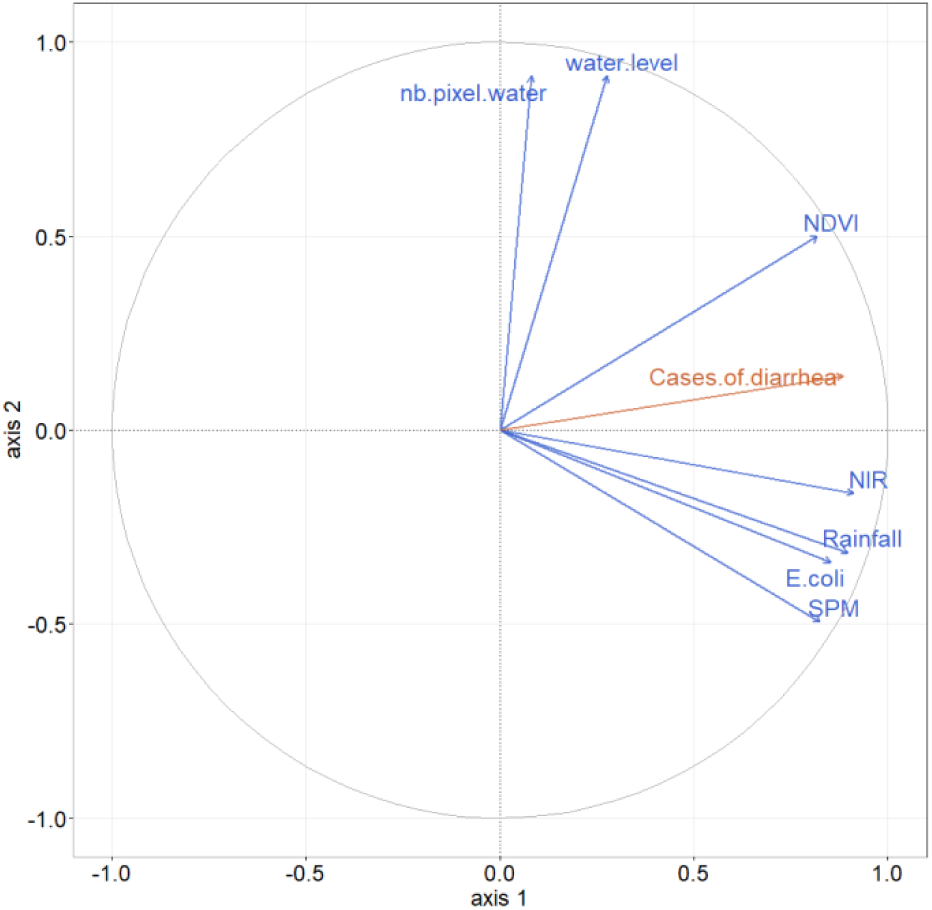
Correlation circle of partial least squares (PLS) regression, illustrating relationships between predictor variables (blue), cases of diarrhea (orange) and the two first PLS components (first component on the x-axis, second one on the y-axis).

**Fig 13.**
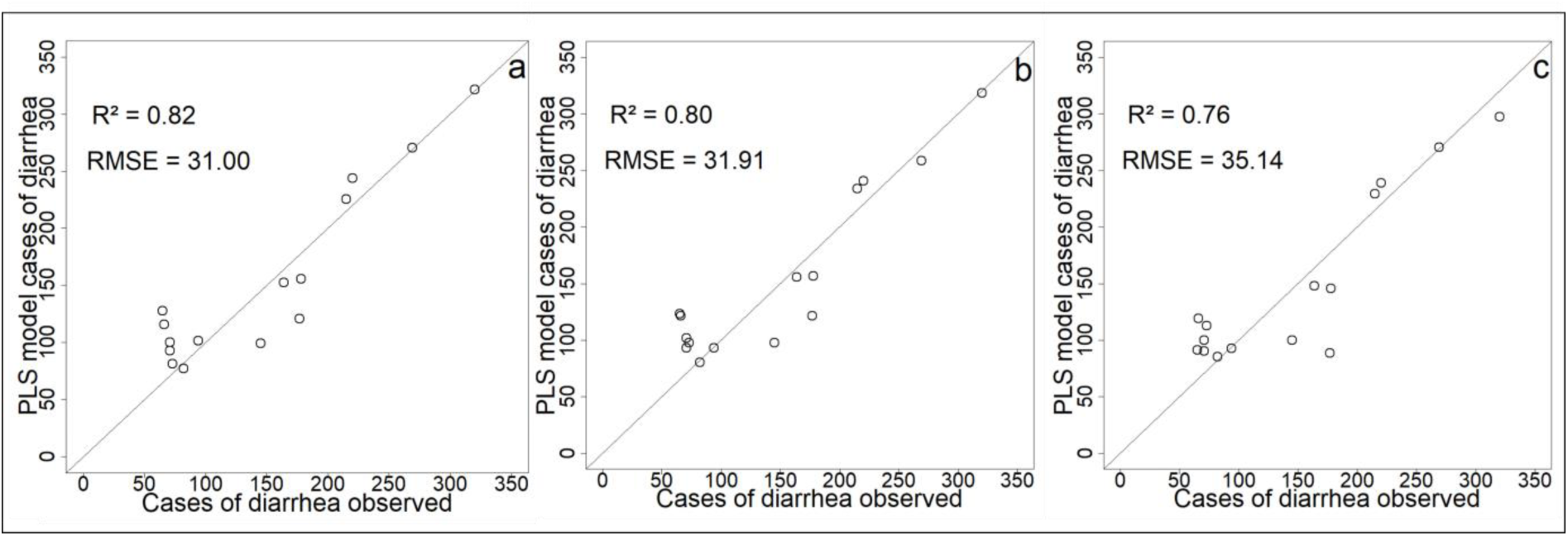
Comparison of models’ predictions with cases of diarrhea observed (a) PLS model with all environmental variables; (b) PLS model with Monthly rainfall, SPM, *E. coli*, NIR Band, and NDVI; (c) PLS model with NIR band, NDVI and Monthly rainfall.

**Fig 14.**
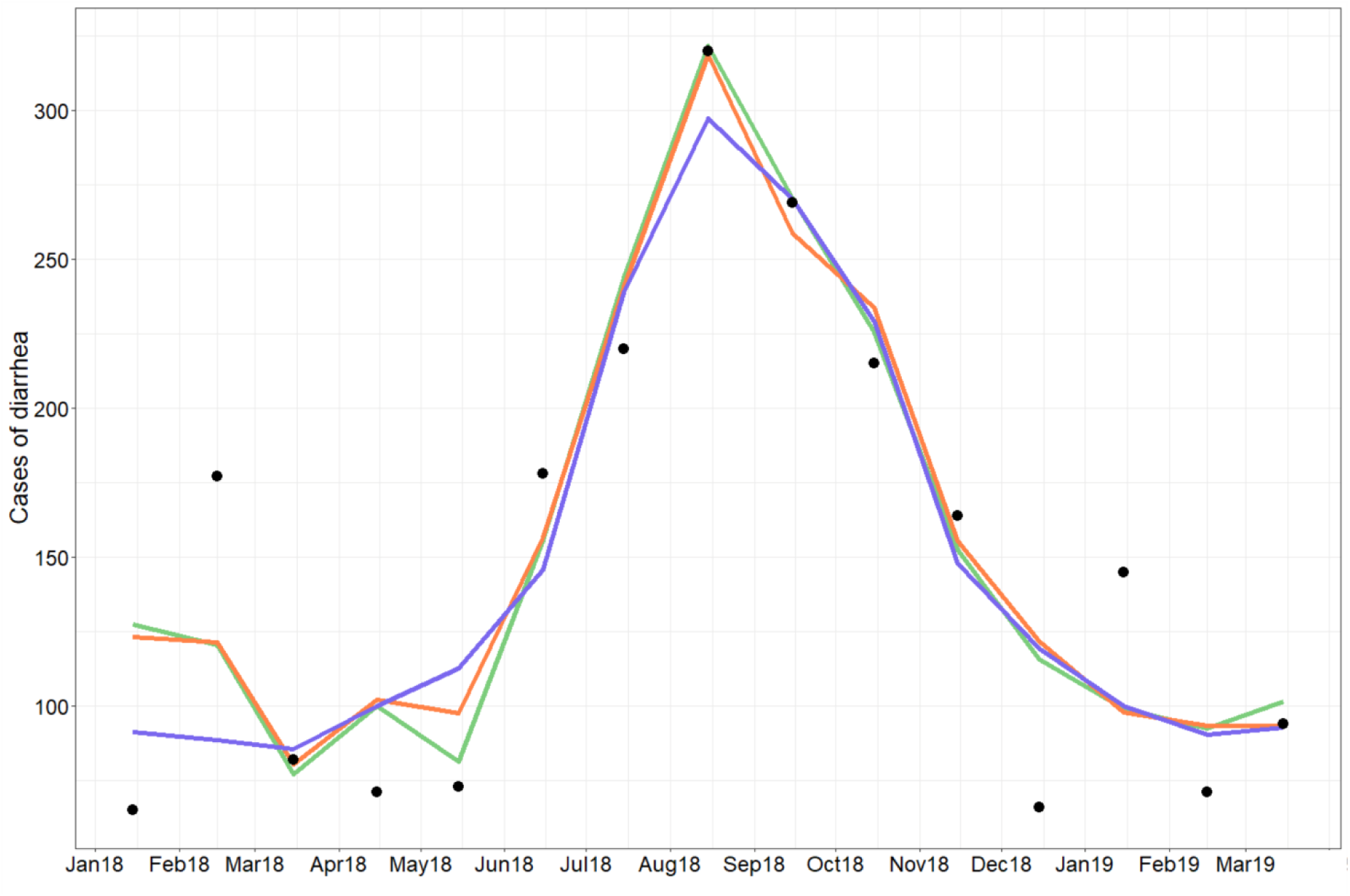
Observed diarrhea (black point) and predicted diarrhea cases using all variables (green line), NIR band, SPM, E.coli and rainfall variables (orange line) and only satellite variables (blue line)

Cases of diarrhea = 1.63 + 0.163 · Monthly rainfall + 0.013 · NIR band + 400.232 · NDVI (2) where Monthly Rainfall is expressed in mm, NIR band in sr^-1^*10000, and NDVI and Cases of diarrhea are unitless.

### Socio-health vulnerabilities

The interviews revealed the different types of water supply used for domestic purposes: borehole, “puisard”, lake and / or tributary, and combinations of these different types of water. People, especially women, often need to draw water 2-3 times a day to fulfil the requirements of the household.

#### Environmental and spatial vulnerabilities

The first criterion of vulnerability is environmental. The evolution of the vegetation cover during the year being similar for the 3 health areas, the environmental vulnerability is mainly linked to the direct proximity to surface water, including Bagre Lake. Thus the higher number of cases of diarrhea recorded at the Dierma health center could be linked to the fact that this health area has the most inhabited areas (Dierma village, Zella, Kapore Peul) in close proximity to surface water (Fig 1). The population would therefore be in contact with surface water (microbiological hazard) all year round, and particularly so in July and August. This situation concerns also Fulani settlements near Lengha (Ouazi Peul, Yakala and Yakala Peul). This vulnerability linked to the proximity to surface water, increases with distance from drinking water points and with the use of “puisard” for part of the year. This mainly concerns the Fulani settlements, in particular Ouazi Peul, as well as certain sectors of Kapore Peul and Yakala Peul, located far from boreholes or wells. "*I use the water from the lake when I don’t have time to go to the borehole*", explained a woman working on a vegetable field (Kapore Peul), and “*the majority goes to the lake or dig ’puisards’”* at Ouazi Peul. This situation is aggravated during the rainy season when access to a borehole becomes difficult: the tracks are less passable, and rivers are full of water, forcing people to bypass them. Moreover, temporary streams and ponds located in proximity to the habitations lead to direct use of surface water, or to digging of “puisards”, thus increasing the use of this type of non-potable water at this time of year. During a so-called “normal rainfall year”, the populations living far from boreholes mainly use the lake between December (the backwaters and tributaries are dry from December-January) and June. Both “puisards” and the lake or the tributaries close to the habitations are used in July and November, with mainly the “puisards” being used between August and October. “Puisards” are often dug close to where people live. The water quality is considered to be better than that of the lake: "*puisards make people less sick*" (B.A, Yakala Peul), "*water from the puisards is better than that of the lake*" (D.M, Ouazi Peul). Indeed, the *E. coli* values observed in the “puisards” are lower: at Kapore “puisard” the values are 480 MPN 100 mL^-1^ without rain and 3 300 MPN 100 mL^-1^ after rain; in comparison, the values on the Kapore routine site in the Bagre lake at the same dates were equal to 5300 and 15000 MPN 100 mL^-1^. Nevertheless, *E. coli* numbers are still high for a water source used for drinking water.

Distance also plays a role in vulnerability in terms of access to healthcare. The Lengha HSPC action plan in 2018 reveals that 41.5% of the population resides between 5 and 10 km, including some populations of Ouazi and Zamsé Peul, and 8% at more than 10 km, mainly Yakala Peul, which limits access to healthcare. Moreover, these difficulties in accessing healthcare are accentuated during the rainy season. The data collected from the action plans of the HSPC in Dierma reveal that from July to December 37.11% of the population had difficulty in accessing the HSPC due to the river that crosses the village (Fig 1B).

This form of vulnerability concerns Yakala Peul, Ouazi Peul, Kapore Peul, especially those living far from potable water points, and to a lesser extent Zamsé Peul and the Zella district of Dierma.

#### Social vulnerability

The lack of financial resources is also a vulnerability criterion for access to water and healthcare. Access to drinking water may be subject to charging: in Dierma it is 500 FCFA / woman / concession, and in Zamsé Peul it is 5000 FCFA / year. The lack of financial resources can therefore lead some people to use the water from the lake: "*this is why we are going to draw water from the lake*" (B.D). The issue of the cost of care, between 1500 and 5000 FCFA, can also be a limiting factor. Financial difficulties can lead to self-medication or the late arrival of some people at the HSPC with conditions "*often more serious and complicated to treat*" (nurse of the HSPC in Lengha). The possibility that the HSPC accepts or refuses credit varies between respondents. Financial availability varies according to the type of activities and the harvest sales periods: for example, for those who practice market gardening there is more money available in November-December and in February-May. Finally, activity type can also be a criterion of socio-health vulnerability. Pastoral activity can limit access to potable water. Similarly, some farmers cultivate fields far from their concession, forcing them to use surface water like those in the Banoma district of Dierma.

Access to water and healthcare also depends on social and political relations inherited from recent local history, in particular that linked to land use policies as well as rights on the various resources (water, land) and associated conflicts. In our study region, the pastoral populations remain particularly vulnerable in terms of access to drinking water and healthcare. For instance, Fulani herders and farmers are subject to different public policies: no health center and few drinking water points are available for the Fulani. Recently, 100 concessions of the Fulani of Yakala were displaced because the inhabitants of Lengha needed more land. The resulting increase in the distance from the Fulani concessions to borehole and the HSPC of Lengha also increased their socio-health vulnerabilities. The Fulani remain subject to subjugation and power relations. This form of vulnerability therefore concerns mainly the Fulani settlements, people cultivating distant fields and the poorest section of the population.

In summary, the epidemiological study and the interviews revealed five criteria of vulnerability: 1) proximity to surface waters and the distance to a potable water point and health center; 2) financial difficulties; 3) age: children who are more fragile and who drink surface water while playing; 4) type of activities: shepherds who leave their habitation with drinkable water and then drink water from the lake, and some people who cultivate on fields far from concessions; 5) population in a situation of subjugation.

### The microbiological health risk and its particularity in 2018

The microbiological health risk associated with diarrheal diseases is higher during the rainy season. The values of *E. coli* are the highest, the use of surface water is more important (closer and easier to access relative to the pumps for people living far from the villages), access to health centers is more difficult (impassable roads, less financial availability). This risk is reinforced for the Fulani by the territorial organization and the associated power relations.

The number of cases of diarrhea was higher in 2018 suggesting that the risk was even higher that year. The low rainfall recorded in 2017 resulted in an exceptional decrease in surface water during the following dry season: water level was 228.16 m on 18 July 2018, compared to 230.154 m on 16 July 2017 and 231.456 m on 20 July 2019). Water level returned to normal at the end of July (230.184 m on 28 July 2018 compared to 230.519 m on 26 July 2017 and 232.736 on 30 July 30 2019). The scarcity of water at the end of the dry season probably has also negatively impacted the water level of the aquifers provoking a reduction in water flows from the borehole and the breakdown of the pumps. Populations usually using borehole or pump water may have had to use water from the lake or dig “puisards” near the lake. Difficulty to access to drinking water and to dig ‘puisards” coupled to environmental parameters could explain the exceptional number of diarrhea cases observed in 2018, with the exception of Dierma.

## Discussion

### Dynamics of *E. coli* in relation to environmental parameters

We have shown that water at the Kapore site is polluted by bacteria of fecal origin throughout the year, with higher values observed during the rainy season. Our *E. coli* numbers are within the range of values found by Akrong et al. [86] in the Bosomtwe Lake in Ghana (*E. coli* ranging from <1 to 5400 cfu / 100ml and for enterococci from <1 to 8400) and by Tyner et al. [87] in Lake Malawi (*E. coli* range from 0 to 21200 cfu / 100 ml).

We also observed a significant relationship between *E. coli* and enterococci, with some variations with time. This relation is in line with data from the Democratic Republic of Congo (R² = 0.94, [88]) and Hawaii (R²=0.72-0.87, [89]). However, we also observed some outliers for two groups of data points. The first group, characterized by high *E.coli* and low enterococci during the period from March to mid-June, may be linked to a phase of resuspension of particle-attached *E. coli* that had previously sedimented, potentially during periods of disturbance, such as when animal are drinking in the lake. The second group (mid-September and late November-early December) is characterised by low *E. coli* and high enterococci. Higher values of enterococci are often associated with fresh fecal matter as enterococci are considered to have more restrictive nutrient requirements than *E. coli* and to have a lower capacity of survival in the environment [90–92]. It is therefore probable that this second group of points corresponds to the presence of recent inputs of fecal matter. Recent work from Alberta, Canada [93] also noted that enterococci were not suitable for the assessment of irrigation water quality. Given the overall good relationship between *E. coli* and enterococci and the questions on the use of enterococci as an indicator of fecal contamination in tropical systems, we have focused the analysis and the following discussion on the relationship between *E. coli* and the environmental parameters. Nevertheless, a more in depth investigation on the use of enterococci as a FIB in tropical surface waters for the evaluation of the risk to human health would not be a miss.

We observed significant positive correlations between *E. coli* and precipitation and in situ SPMs. High intensities of convective rainfall in this area, when vegetation is not yet fully developed and when erodible material is present, create surface runoff that is likely to transport SPM as well as bacteria towards the Bagre Reservoir. Other studies [94, 39, 95] have also observed positive correlations between *E. coli* concentrations and precipitation and suggested that this relationship was due to contamination of the soil surface by bacteria of fecal origin from animal husbandry and open defecation. Indeed, in rural areas in Burkina Faso in 2017, 63% of people were supposed to be concerned by open defecation due to the lack of latrines [5]. During precipitation events, this matter is transported via runoff to downstream water bodies. Therefore, this type of diffuse source is often a more important source of fecal contamination during the rainy season than during the dry season. Finally, in addition to being transported down slopes, as is the case for SPM, *E. coli* could also have local point sources (livestock, open defecation near the routine point) which may explain some of the high values observed (for example at the beginning and at the end of August).

The link between precipitation, land use and erosion, SPM and *E. co*li concentrations has been observed elsewhere. In tropical environments, Nakhle et al. [96] in the Mekong basin in Lao PDR observed that concentrations of *E. coli* are positively correlated with total suspended sediment (TSS) concentrations, particularly in watersheds subject to soil erosion. Petersen and Hubbart [97] in an Appalachian watershed of West Virginia (USA) found a strong positive relationship between SPM and *E. coli* that was strongly related to the percentage of agricultural land. A similar relationship between land use, SPM and *E. coli* was also observed by Tong and Chen [98] working in the State of Ohio. Poor microbial water quality is often associated with high relative percentages of cultivated land in both tropical and temperate regions [99–100]. The watershed of the Bagre Reservoir has undergone profound changes since the 1980s with an increase in cultivated areas and bare soils to the detriment of natural vegetation [67, 68, 101], which may have resulted in increased erosion and particle transport towards the lake. This type of land use can also alter the size of the particles [102] leading to increases in the proportion below 5µm [103]. This smaller fraction is known to be correlated with *E. coli* [97] and can extend the survival of bacteria in organic carbon rich environments [104]. Indeed, particle size distribution, determined from three water samples from the Bagre Reservoir in 2017 using a particle size analyzer Partica LA-950V2, has a bimodal structure with very fine particles (0.2-0.5µm and 1.2-13.2µm) which may have exerted an impact on the presence and survival of *E. coli*. More generally, the presence of these very fine particles is probably linked to characteristics of the soil type in this region as they have also been observed in the Agoufou region (Mali) [75].

We observed a lag of about 15 days between the peak of SPM and *E.coli* in the summer. The period (end of July - beginning of August) corresponds to a period of high vegetation cover. Vegetation cover, and particularly riparian vegetation, plays a strong role in filtering overland flow and reducing soil, and probably bacteria, downslope transport [105]. In this context, the time lag between the peaks of SPM and *E. coli* may be explained by the conjunction of several phenomena: 1) the remobilization/resuspension of bacteria in mud during flooding of the river banks (Fig 6E): satellite images show that the main flooding of the banks took place between 11^th^ and 31^st^ July 2018 (Fig 15); and 2) the presence of ideal conditions for the development of bacteria. Indeed, as the sampling point was close to the banks, bacteria at this time of the year can be considered to be in an environment conducive to their development: protection against UV and support by the presence of SPM, but also presence of sediment/sludge rich in carbon, and nutrients. SPM is known to increase the availability of nutrients and organic matter and provides some protection from light exposure [106–107] and *E.coli* often have higher growth and survival rates when associated with SPM [22, 108]. Thus, the flooding of the banks with the remobilization of bacteria present in the mud, the increase in the survival rate, and their higher relative proportion in the attached fraction (and as such protected from light exposure by the presence of SPM) may well explain this time lag between the peaks of SPM and *E. coli*.

**Fig 15.**
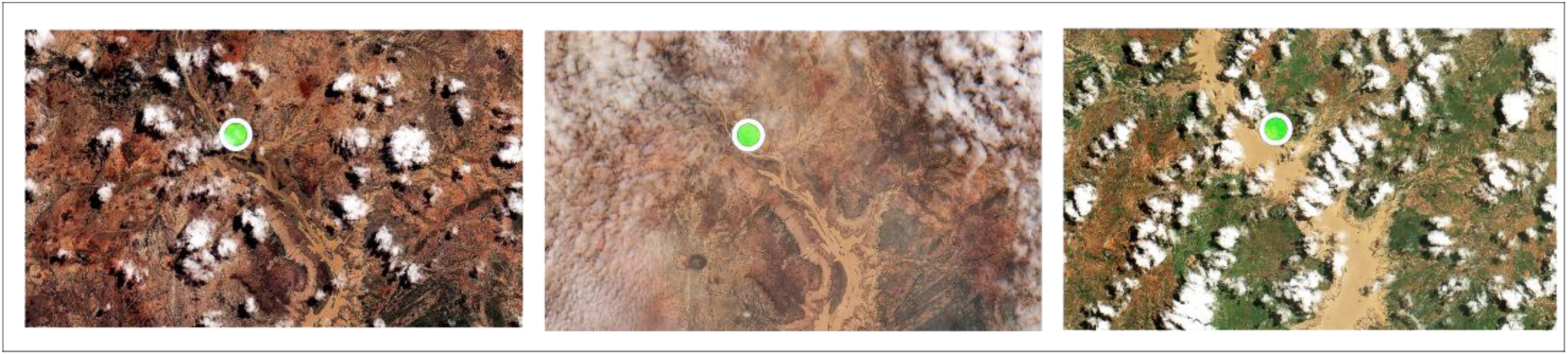
Evolution of the surface water area in the region of the routine measurement during the month of July 2018 (from left to right: Sentinel-2 images from 01/07/2018, 11/07/2018 and 31/07/2018)

Ultimately, the use of satellite data represents a major challenge with a very cost-effective way of predicting the dynamics of *E. coli* via the characterization and monitoring of environmental determinants such as SPM, land-use, and precipitation. Previous work has shown the importance of using landscape data from satellite images to characterize water quality, such as Johnson et al. [109] and Hua [110], and to monitor SPM dynamics [75, 78].

### Relationship between diarrhea incidence, environmental variables and *E. coli*

Diarrhea case incidence shows a marked seasonality with a main peak during the rainy season and a second during the dry season. Such positive associations have been also observed between cases of diarrhea and rainfall in Ghana [111] and during the peak monsoon rainfall months in the Philippines [112], Bangladesh [113] and Lao PDR [26]. In addition, we show that the increase in *E. coli* is associated with an increase in cases of diarrhea similar to what has previously been observed in a river in Botswana [114]. We did not find any link with *E. coli* or the environmental parameters for the second peak in January-February. Nitiema et al. [115] as well as Bonkoungou et al. [116] found a stronger presence of rotavirus between December and February-March in Burkina Faso and Mwenda et al. [117] also report that, in several African countries, the prevalence of rotavirus peaked during the cool, dry months. Therefore, it is possible that rotavirus, and not *E. coli*, was responsible for the secondary peak in diarrhea cases. Moreover, the NIR band reflectance data miss the peak of diarrhea linked to rotavirus. If the cooler winter months (December to February) are excluded, the monthly cases of diarrhea are strongly correlated with the NIR band data (Rs² = 0.83**).

### Predictive models of *E. coli* and cases of diarrhea

Few studies have reported predictive models using independent variables to explain *E. coli* concentration and of these, the majority concern temperate areas e.g. [118–119]. Some recent studies have looked at this question in tropical areas of Asia [120, 96]. Common with our study, these other works use turbidity or/and Total Suspended Solid (TSS) associated with other environmental variables. The model suggested by Chen and Chang [118] includes antecedent precipitation, stream temperature, and TSS concentration, and gives an R² of 0.60 for the Johnson catchment, which is moderately urbanized. Hamilton and Luffman [119] used a multiple regression with an R² of 0.57 to predict *E. coli* in the Little River (Tennessee, USA) using antecedent precipitation, discharge and turbidity. Boithias et al. [120] used also a model to predict *E. coli* concentrations including TSS and discharge with an R² of 35.3%. Nakhle et al. [96] used PLS regression and highlighted that *E. coli* was mainly explained by areal percentages of unstocked forest, turbidity and TSS. Our model retaining the most significant variables (NIR, precipitation, SPM) and the one based on satellite data (NIR and precipitation) present similar results with an R² = 0.54 and an R² = 0.48 respectively (without individuals below the detection threshold of *E. coli*: R² = 71 and R² = 60 respectively). In future research, it would be interesting to also integrate water temperature and discharge into the regression.

There are many studies that highlight the link between diarrhea incidence and meteorological parameters such as rainfall [52, 43–44, 121]. More specifically in Sub-Saharan Africa, both Alexander et al [114] in Bostwana, and Alemayehu et al [122] in Ethiopia highlighted this relationship. However, to our knowledge, the study presented here is the first to illustrate the link between diarrhea incidence and satellite-derived variables. Some explaining variables probably have causal links with diarrhea cases (*E. coli* and rainfall for instance), while others may only show spurious correlation (NDVI probably). At this stage, the monthly time-step and the well-marked seasonal cycle of different variables prevents more conclusions to be drawn. A possible time-lag between SPM, NIR and cases of diarrhea cases may impact the correlation value given. The number of data is too small to give solid evidences so far, although such a lag is expected because of delays in contaminants peak, contamination, and consultations in health center.

The validity of these models over several years and their capability to predict interannual variability will have to be assessed in further studies.

### Epidemiology and socio-health vulnerabilities

In Dierma and Dango, consultations are more frequent in the dry season than in the rainy season. This is probably linked to the agricultural calendar and to the fact that the inhabitants of Dierma, and to a lesser extent of Dango, cultivate crops on the other side of the lake. Ouedraogo and Janin [66] also observed seasonal differences in the use of healthcare for all pathologies in the region studied.

Our interviews highlight a more important use of surface water during the rainy season. This is due to the easier accessibility and availability of surface water as opposed to borehole water, similar to the conclusions of Schweitzer et al. [64] for Burkina Faso. In future research, it would be interesting to determine the proportion of people using the different types of water (lake, other surface water, “puisard”) and to investigate the seasonality of FIB and SPM across a wider range of water surface points including a more comprehensive investigation of the “puisards”.

Distance and financial availability can be limits for seeking healthcare. This is in line with results by Haddad et al. [63], who have shown that people living more than 10km away from a health center went less to health facilities: 25% went there on the 7th day of the illness against 40% for those living at less than 10km. Haddad et al. [63] also revealed that the maximum cost envisaged to pay for treatment for a family member is 2537 FCFA in rural areas. However, in our study region, the cost varies between 1500 and 5000 FCFA, which can further limit the use of care. Finally, our interviews revealed that financial availability for treatment varies according to the harvest sales periods in agreement with Schweitzer et al. [64].

Social, physical and financial inequalities, problems of marginality and power imbalances also play an important role in determining access to water resources and health care [123–125]. They are therefore important determinants of socio-health vulnerabilities. Regarding access to water, this refers to "political ecology" which "*recognizes that access and control over resources, including water, are rooted in local stories and social relations*" [126–127]. Indeed, our interviews reveal that the notions of territory, marginality, and social exclusion shape the landscape of water resources. This theoretical framework can also be extended to include access to care. Thus, social and political interactions between resource managers and health actors on the one hand and potential users on the other hand are often very limited. This is particularly the case for marginal populations such as the Fulani [128, 7, 129]. These socially constructed and politically imposed inequalities lead to an unequal distribution of access and use of water and health care, as Dinko et al. [127] has shown in Ghana.

Ultimately, the socio-health vulnerabilities and therefore the associated health risks observed in our study region combine social, political and physical factors. The study of the health risk linked to diarrheal diseases requires an interdisciplinary analysis using both qualitative and quantitative methods. However, these approaches are still rare as already pointed out by Batterman et al. [130].

Finally, we would like to underline the limits of this case study. Our study concerns a small region (120 km²) with a single in-situ monitoring point over one year and some occasional measurements in “puisard”. We have only monthly data on diarrhea cases from 3 health centers, and interviews with inhabitants of the 3 villages, Fulani settlements in the area and health personnel. It would be interesting to have a longer time series of data and to conduct a quantitative vulnerability survey. Despite these limitations, our work allowed to 1) test hypotheses on the link between *E. coli*, environmental variables and cases of diarrhea in a tropical context, 2) develop a simple prediction model for *E. coli* from variables that can be estimated using satellite data; 3) propose a first step in the interdisciplinary analysis of the health risk by combining environmental, microbiological, epidemiological data and interviews.

## Conclusion

This study highlighted the positive correlation between *E. coli* and enterococci in tropical rural West Africa with higher numbers observed during the rainy season. *E. coli* and diarrheal diseases were strongly correlated with precipitation, in-situ SPM, and reflectance in the NIR band between February and mid-December implying that satellite data can provide an important input to water quality monitoring. Such a tele-epidemiological approach that incorporates environmental, microbiological and epidemiological variables is of particular interest in regions where in-situ measurements are absent or scarce and where the impacts of global and local change (land use change, modifications in precipitation) are most strongly felt.

In the study area, diarrheal diseases represent the 3rd cause of consultation, with children under 5 years old the most impacted. The vulnerability of the population depends on income, age, activity, and worsens during the rainy season due to reduced accessibility to drinking water and healthcare in conjunction with increased proximity of water sources of poor quality. The importance of the power relations, which have a direct consequence on the territorial organization for access to water and healthcare, were also highlighted. Overall, the microbiological health risk associated with diarrheal diseases is more important during the rainy season from June to September. Therefore intervention though public education/awareness should be increased as the rainy season approaches.

Cross-referencing of data from interviews and environmental parameters helped to clarify the causes of the large number of cases of diarrhea in 2018, illustrating the importance of interdisciplinary approaches. In order to have a more detailed understanding of the processes and interactions at play, it will be necessary in the future to investigate the co-variability of environmental, microbiological and epidemiological data over finer timescales and longer periods. This should be coupled with a more in depth quantitative survey (large number of interviews) to obtain a complete vision of health issues related to fecal contamination over the whole Bagre area through an EcoHealth approach.

Although this is an initial case study, this work provides is a first step in the interdisciplinary analysis of health risk. The approach proposed here provides a base for developing future programs that explicitly integrate global, social and human mechanisms that modify habitats, modes of transmission, pathogen survival, and ecosystem functioning, as well as access to drinking water and healthcare. It is only by explicitly recognizing the complexity of the problem and by adopting a more equitable approach to problem solving that we can hope to provide solutions and to improve the livelihoods of the affected populations in Burkina Faso and elsewhere.

## Data Availability

The in-situ data file is available from a confidential link on the dataverse website
After publication, the link will be public.

## Acknowledgements

The authors thank Ali Niaoné for the field measurements, Marielle Gosset for advice on precipitation datasets, Bruno Lartiges for discussion on bacteria-particle characteristics, Kpeke Nestor Kambiré and Yasmina Karambiri for collecting part of the health data and part of the location of village concessions, boreholes and wells, and the regional health office of Tenkodogo for the use of its premises.

## Footnotes

1 The official Joint Monitoring Program definition is the percentage of the population with access to at least 20 l of water per person per day from an improved drinking water source (household connections, public standpipes, boreholes, protected dug wells, protected springs, and rainwater collection) within one kilometre of the user’s dwelling.

