## Supplementary material for "Environmental determinants of *E. coli*, link with the diarrheal diseases, and indication of vulnerability criteria in tropical area (Kapore, Burkina Faso)": S1 Fig, S2 Fig, S3 Fig, S4 Fig, S5 Fig, S1 Table, S2 Table

### Supporting Information

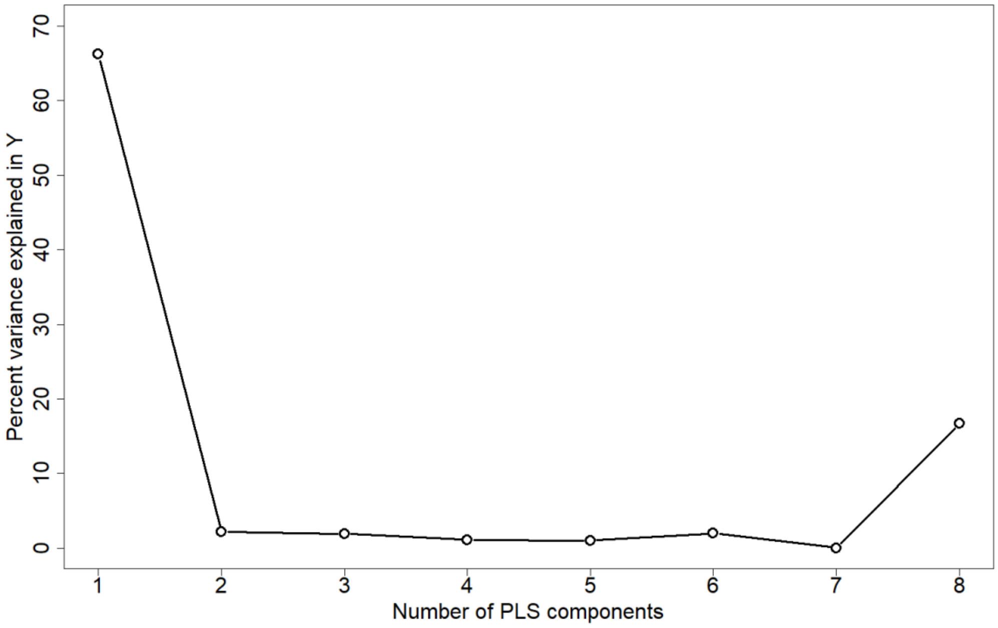


**S1 Fig. Percent variance of *E. coli* from environmental variables for each component**

| Variable | Percent |
|---|---|
| SPM | 79 |
| Weekly Rainfall | 71 |
| Water level | 4 |
| Cumulative rainfall | 9 |
| Nb pixel water | 3 |
| NDVI | 32 |
| NIR band | 83 |
| Daily rainfall | 37 |

**S1 Table. Percentage of the variability of *E. coli* explained by each of the variables for the 1st component**

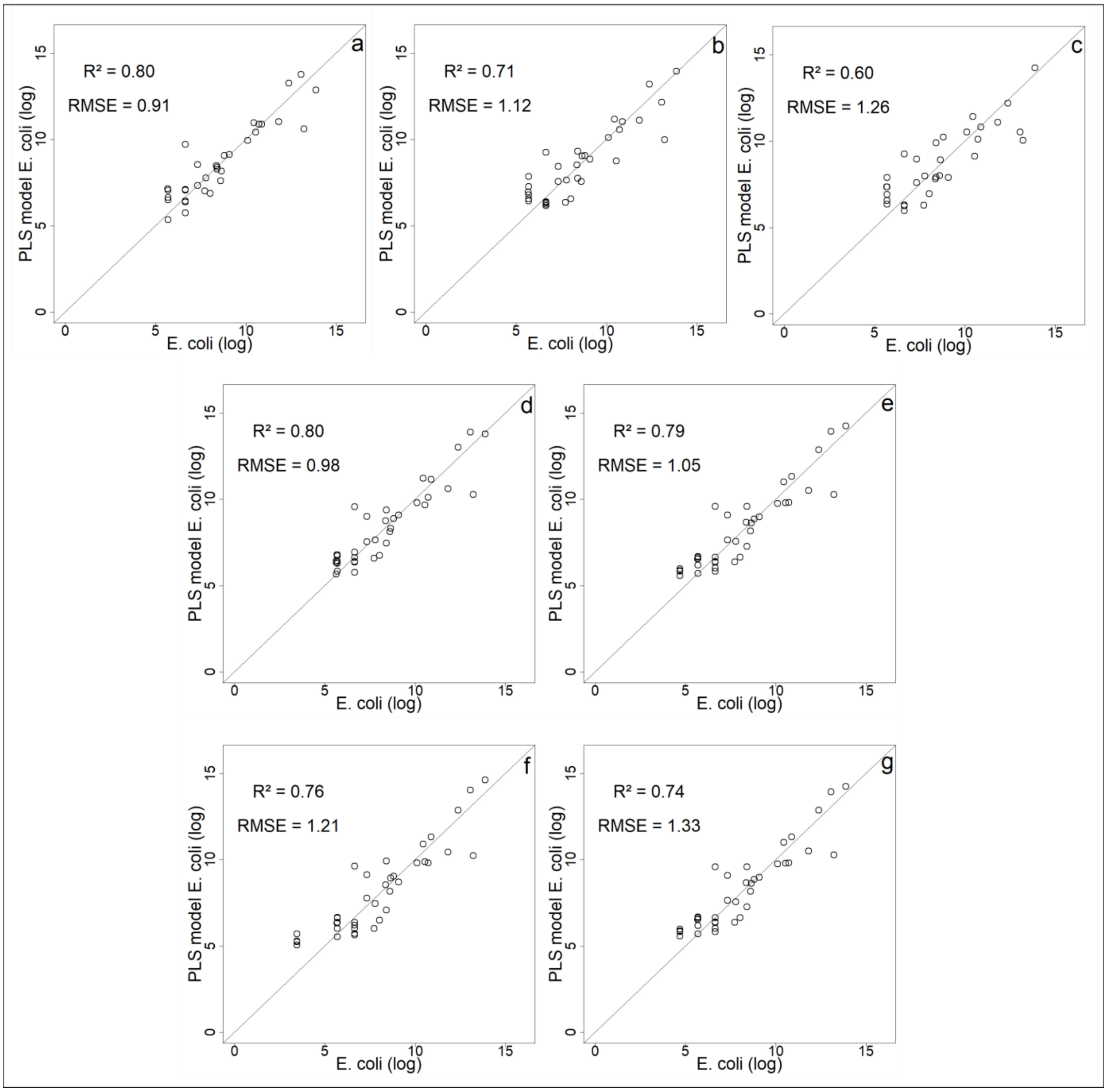


**S2 Fig. Comparison of models' predictions with observed *E. coli* (log values) (a) PLS model with all environmental variables without individuals 31-34; (b) PLS model with NIR band, SPM and Weekly rainfall without individuals 31-34; (c) PLS model with satellite variables only (NIR band and Weekly rainfall) without individuals 31-34; (d) PLS model with all environmental variables and value 49 assigned for the values less than 50 MPN 100mL$^{-1}$; (e) PLS model with all environmental variables and value 25 assigned for the values less than 50 MPN 100mL$^{-1}$; (f) PLS model with all environmental variables and value 10 assigned for the values less than 50 MPN 100mL$^{-1}$; (g) PLS model with all environmental variables and value 5 assigned for the values less than 50 MPN 100mL$^{-1}$**

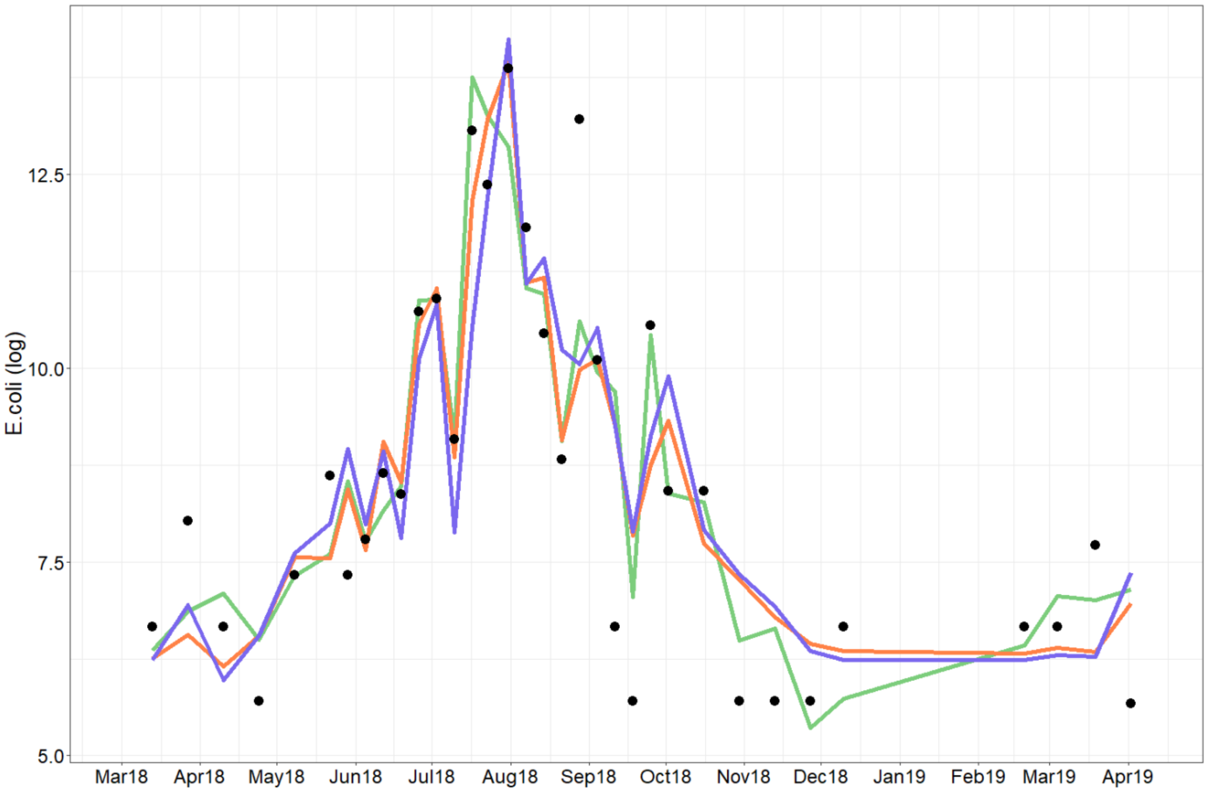


**S3 Fig. Observed *E. coli* (black point) and predicted *E. coli* (a) without individuals 31-34 (black line for all variables, red line for NIR, SPM and Rainfall variables, blue line for satellite variables**

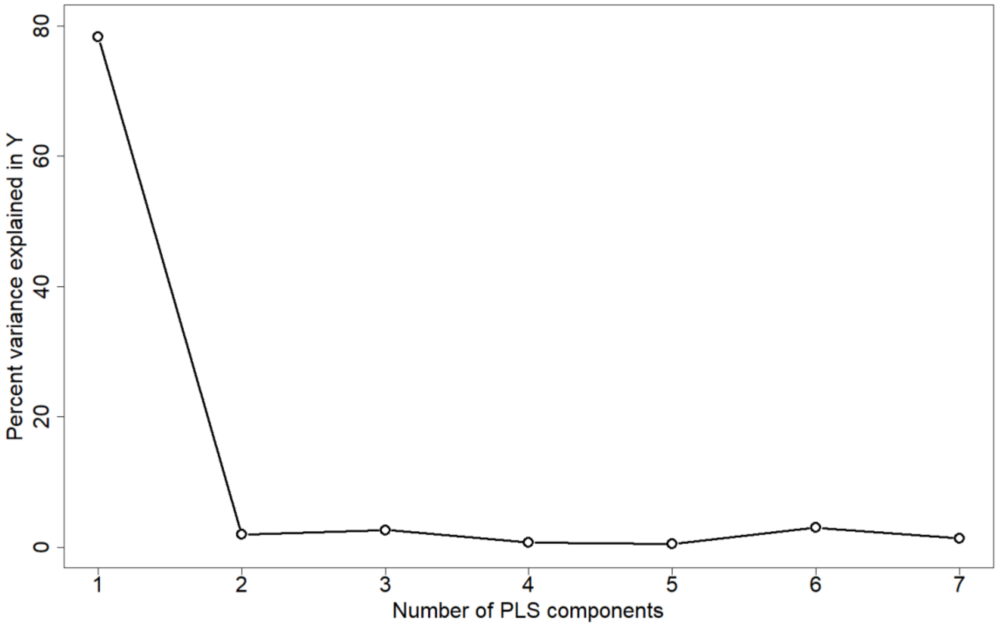


**S4 Fig. Percent variance of cases of diarrhea from *E. coli* and environmental variables for each component**

| Variable | Percent |
|---|---|
| **SPM** | **68** |
| **Rainfall** | **80** |
| **Water level** | **4** |
| ***E. coli*** | **73** |
| **Nb pixel water** | **0.6** |
| **NDVI** | **67** |
| **NIR** | **83** |

**S2 Table. Percentage of the variability of cases of diarrhea explained by each of the variables for the 1st component**

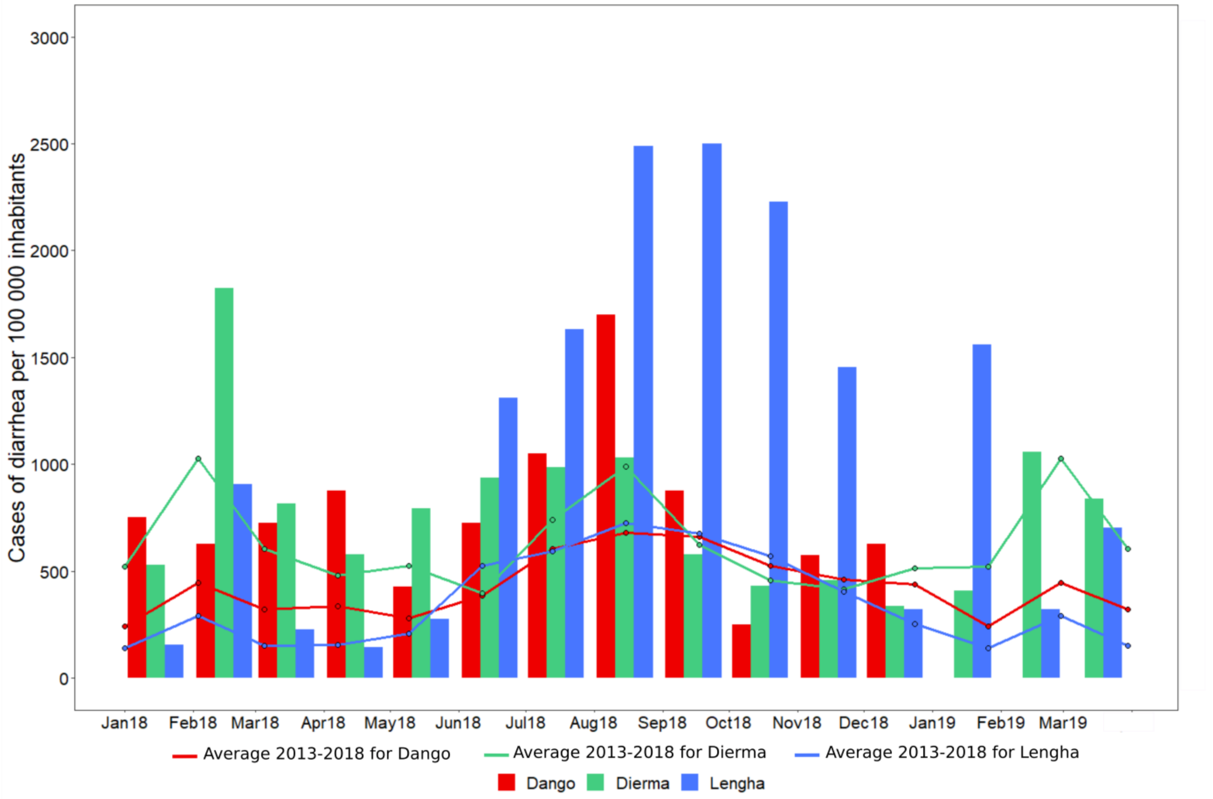


**S5 Fig. The incidence rate from diarrheal diseases per 100 000 inhabitants according to the population of each health area**
